# Automation of Systematic Reviews with Large Language Models

**DOI:** 10.1101/2025.06.13.25329541

**Authors:** Christian Cao, Rohit Arora, Paul Cento, Adil Budak, Katherine Manta, Elina Farahani, Matthew Cecere, Anabel Selemon, Jason Sang, Ling Xi Gong, Robert Kloosterman, Scott Jiang, Richard Saleh, Denis Margalik, James Lin, Jane Jomy, Jerry Xie, David Chen, Jaswanth Gorla, Sylvia Lee, Kelvin Zhang, Jennifer Kuang, Harriet Ware, Mairead Whelan, Bijan Teja, Alexander A. Leung, Rahul K. Arora, Jennifer Pillay, Lisa Hartling, Michael Noetel, David B. Emerson, Allan S. Detsky, Andrea C. Tricco, George M. Church, David Moher, Niklas Bobrovitz

## Abstract

**Importance:** Systematic reviews (SRs) inform evidence-based decision making. Yet, many take over a year to complete, are labor intensive, prone to human error, and face reproducibility challenges; thus limiting access to timely and reliable information.

**Objective:** To validate a large language model (LLM)-based workflow (otto-SR) to automate three of the most labour intensive tasks in performing SR’s: article screening, data extraction, and risk of bias assessment; and to assess its feasibility in rapidly updating existing reviews.

**Design, setting, and participants:** We conducted a validation study in four phases, with direct benchmarking against graduate-level human researchers in phases 1 and 2. Phase 1: article screening performance was measured across 32,357 citations from 5 systematic reviews. The reference standard consisted of the original reviews’ screening decisions after full-text screening. Phase 2: data extraction performance was measured across 4,495 data points from 495 studies in 7 reviews. Phase 3: risk of bias assessment (ROB2, Newcastle-Ottawa, QUADAS2) performance was measured across 345 studies from 12 reviews. Reference standards for Phase 2 and Phase 3 were created after blinded adjudication of the original review extraction and RoB assessments. Phase 4: otto-SR was used to reproduce and update the primary analysis from an issue of Cochrane reviews (n=12 reviews, 146,276 citations), with analytical comparisons to the original meta-analyzed findings. All discrepancies underwent dual human review.

**Results:** *otto-SR* showed high performance in phase 1 article screening (*otto-SR*: 96.7% sensitivity, 97.9% specificity; human: 81.7% sensitivity, 98.1% specificity) and phase 2 data extraction (*otto-SR*: 93.1% accuracy; human: 79.7% accuracy). In phase 3, *otto-SR* demonstrated high interrater reliability for risk of bias judgements (ROB2 0.98, Newcastle-Ottawa 0.95, QUADAS2 0.74; Gwet AC2). In phase 4, *otto-SR*, reproduced and updated the primary analysis from an issue of Cochrane reviews. Across Cochrane reviews, *otto-SR* incorrectly excluded a median of 0 studies (IQR 0 to 0.25), and found nearly twice as many eligible studies compared to the original authors (n= 114 vs. 64). Meta-analyses based on *otto-SR* generated screening and extraction outputs, subsequently verified through dual human review, yielded newly statistically significant effect estimates in 2 reviews and negated significance in 1 review.

**Conclusions and relevance:** LLMs have high performance in article screening, data extraction, and risk of bias assessments. They can rapidly reproduce and update existing systematic reviews, laying the foundation for automated, scalable, and reliable evidence synthesis.

## Introduction

Systematic reviews (SRs) are a key element of evidence-based decision-making.^[1]^ However, SRs are incredibly resource-intensive, and can take over 16 months and cost upwards of $100,000 to complete.^[2,3]^ Delays in completing SRs can have major consequences for patients and members of the public, including failure to adopt effective therapies, or prolonged use of ineffective or harmful practices initially supported by less rigorous evidence.^[4,5]^

While several tools have been developed to accelerate SRs,^[6,7]^ none are capable of full automation with human-level accuracy. However, large language models (LLMs) offer new avenues to progress towards automation with their ability to process and reason about natural language. We previously demonstrated that LLMs can achieve high performance in article screening for systematic reviews.^[8]^ Other recent work has demonstrated promise for LLMs in data extraction,^[9,10]^ and risk of bias assessments.^[11,12]^ However, those studies have used small datasets, or relied on author-generated reference standards.

We introduce an LLM-based workflow (*otto-SR*) to support automated and human-in-the-loop (i.e., semi-automated with human oversight) SR workflows. Our framework uses GPT-4.1 (OpenAI) for screening articles, o3-mini-high (OpenAI) for data extraction, and GPT-5 (OpenAI) for risk of bias assessments. We evaluate our workflow on these core SR components in three phases: (1) article screening, (2) data extraction, and (3) risk of bias assessment, with direct comparisons to traditional human workflows and other SR automation tools. In a fourth phase (4), we assessed real-world utility by reproducing and updating the primary analyses from an issue of Cochrane SRs (n=12 reviews) using *otto-SR*.

## Methods

### An agentic workflow for systematic review automation

SR workflows begin with defining a research question and performing a comprehensive keyword search for relevant citations.^[13]^ These citations undergo abstract and full-text screening by two human reviewers independently, with disagreements resolved by a third reviewer. The final set of relevant articles undergoes data extraction and risk of bias assessment by two human reviewers independently, again adjudicated by a third reviewer. The complete human workflow is illustrated in Figure 1 (top).

**Figure 1.**
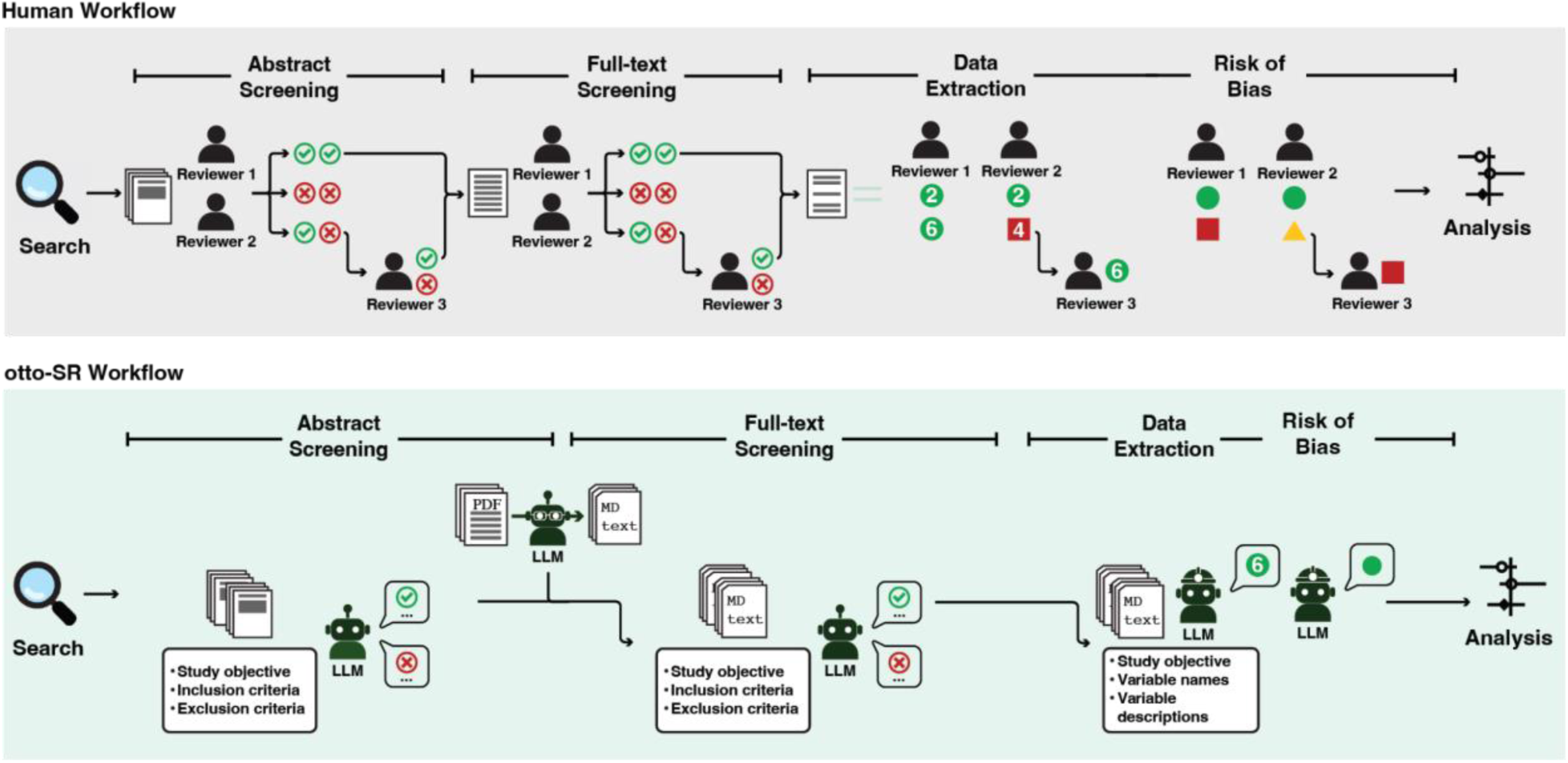
An automated systematic review workflow using LLMs. Infographic displaying the end-to-end SR process for humans (grey) and *otto-SR* (green). Inputs to *otto-SR* include the study protocol (objectives, eligibility criteria), search results, and defined extraction variables. Abbreviations: Markdown file (MD).

*otto-SR* is an end-to-end LLM-based workflow supporting both automated and human-in-the-loop systematic reviews. Citations identified from the original search are uploaded in RIS format to the *otto-SR* screening agent, which uses GPT-4.1 to screen and select for inclusion abstract and full-text articles as a standalone reviewer. The resulting set of included articles is passed to the *otto-SR* extraction agent, which performs data extraction with the o3-mini-high model, and the *otto-SR RoB agent*, which performs risk of bias assessments with the GPT-5 model. For full-text screening and data extraction, retrieved PDFs are processed by Gemini 2.0 flash and converted into structured Markdown (MD) files for downstream tasks. An overview of the *otto-SR* workflow is provided in Figure 1 (bottom).

### Phase 1: Evaluation of Systematic Review Screening Performance

We gathered five SRs via convenience sampling across four types of Oxford Centre for Evidence Based Medicine review questions: SeroTracker dataset^[14]^ (prevalence), the Reinfection^[15]^ and PA-Outcomes^[16]^ datasets (intervention benefits), the PA-Testing^[17]^ dataset (diagnostic test accuracy), and SVCF^[18]^ dataset (prognosis); see Table 1 for review details.

**Table 1.**
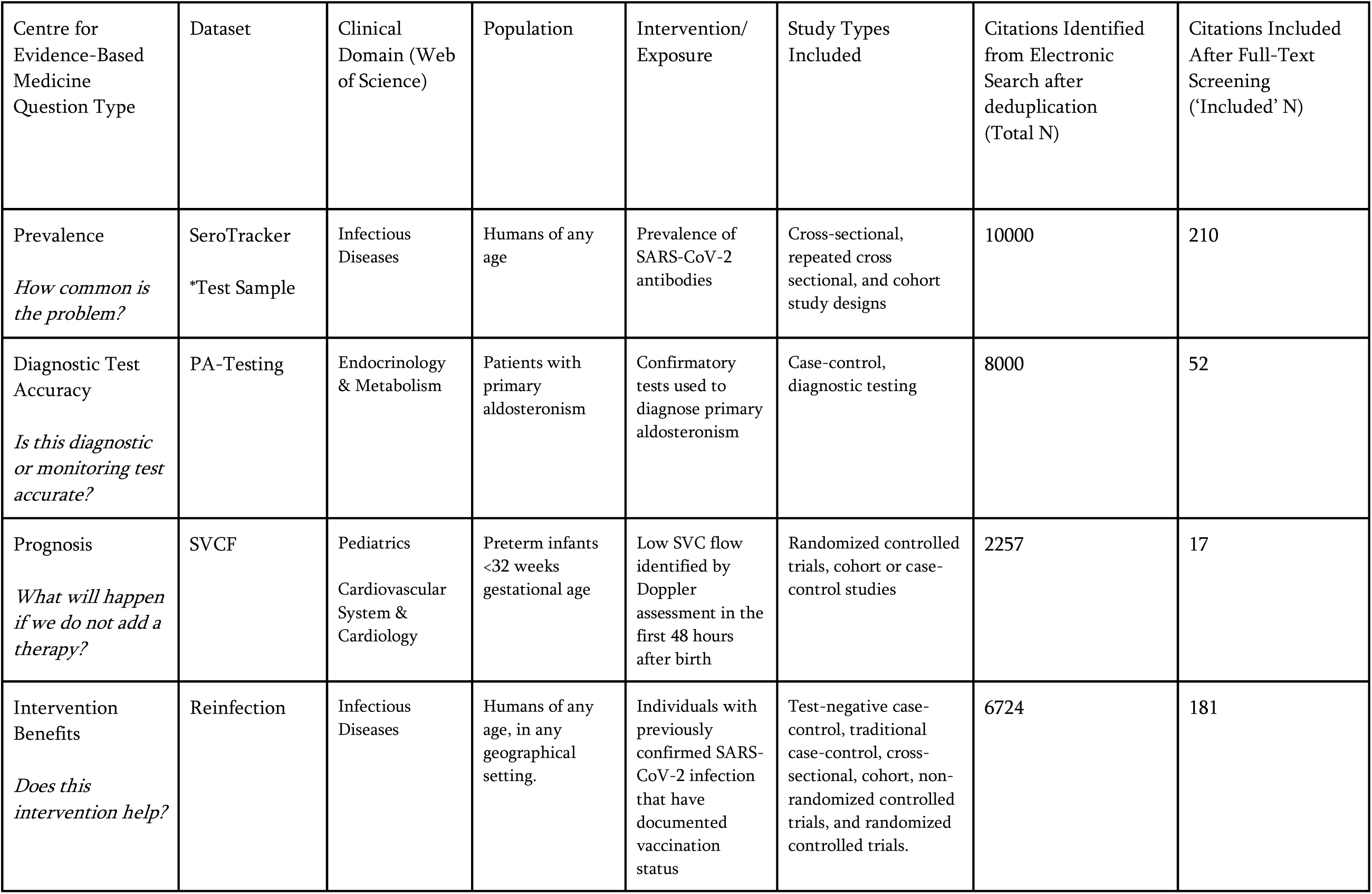
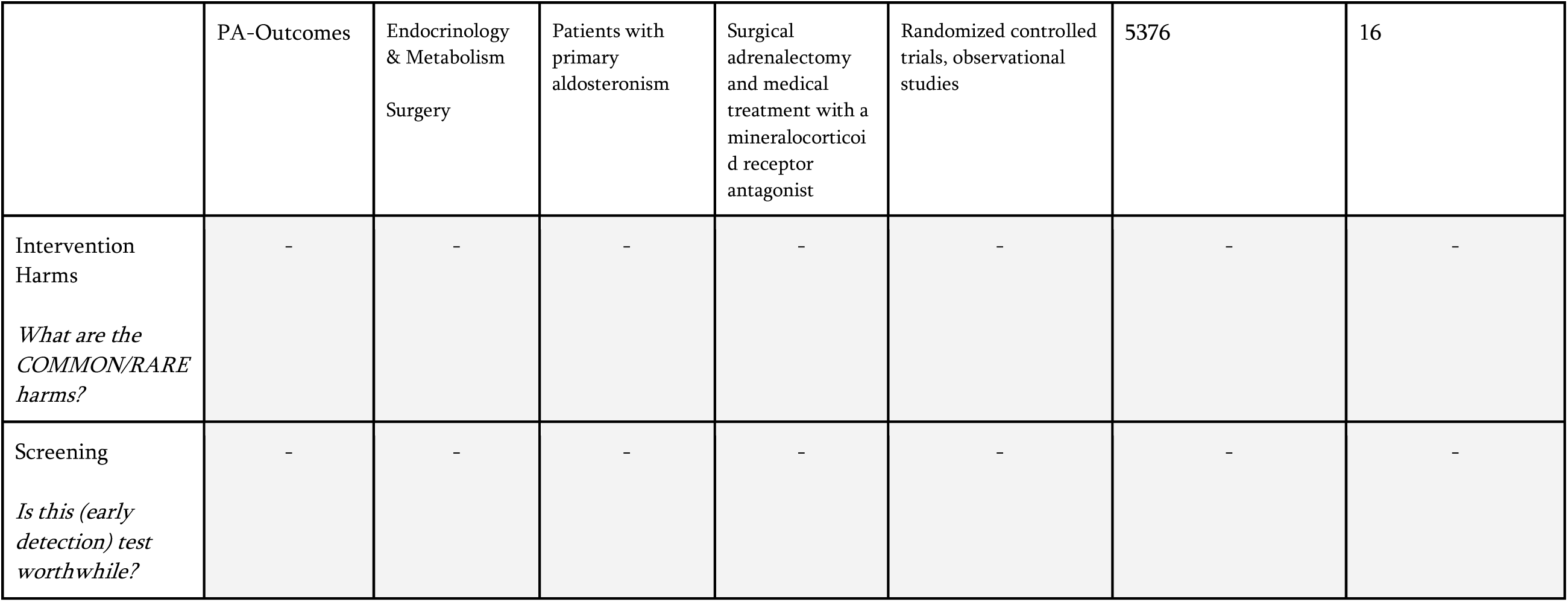
Phase 1: Descriptive overview of datasets used in article screening.

| Centre for Evidence-Based Medicine Question Type | Dataset | Clinical Domain (Web of Science) | Population | Intervention/ Exposure | Study Types Included | Citations Identified from Electronic Search after deduplication (Total N) | Citations Included After Full-Text Screening ('Included' N) |
| --- | --- | --- | --- | --- | --- | --- | --- |
| Prevalence<br>*How common is the problem?* | SeroTracker<br>*Test Sample | Infectious Diseases | Humans of any age | Prevalence of SARS-CoV-2 antibodies | Cross-sectional, repeated cross sectional, and cohort study designs | 10000 | 210 |
| Diagnostic Test Accuracy<br>*Is this diagnostic or monitoring test accurate?* | PA-Testing | Endocrinology & Metabolism | Patients with primary aldosteronism | Confirmatory tests used to diagnose primary aldosteronism | Case-control, diagnostic testing | 8000 | 52 |
| Prognosis<br>*What will happen if we do not add a therapy?* | SVCF | Pediatrics<br>Cardiovascular System & Cardiology | Preterm infants <32 weeks gestational age | Low SVC flow identified by Doppler assessment in the first 48 hours after birth | Randomized controlled trials, cohort or case-control studies | 2257 | 17 |
| Intervention Benefits<br>*Does this intervention help?* | Reinfection | Infectious Diseases | Humans of any age, in any geographical setting. | Individuals with previously confirmed SARS-CoV-2 infection that have documented vaccination status | Test-negative case-control, traditional case-control, cross-sectional, cohort, non-randomized controlled trials, and randomized controlled trials. | 6724 | 181 |
|  | PA-Outcomes | Endocrinology<br>& Metabolism<br>Surgery | Patients with<br>primary<br>aldosteronism | Surgical<br>adrenalectomy<br>and medical<br>treatment with a<br>mineralocorticoid<br>receptor<br>antagonist | Randomized controlled<br>trials, observational<br>studies | 5376 | 16 |
| Intervention<br>Harms<br>*What are the*<br>*COMMON/RARE*<br>*harms?* | - | - | - | - | - | - | - |
| Screening<br>*Is this (early*<br>*detection) test*<br>*worthwhile?* | - | - | - | - | - | - | - |

#### Development of LLM Screening Agent

We developed a novel LLM-based screening agent using GPT-4.1 (OpenAI), adapted from our previously validated *ScreenPrompt* approach,^[8]^ to screen articles at abstract and full-text stages (Supplementary Methods). The agent was prompted using the original, unaltered objectives and eligibility criteria from each respective review protocol (Supplementary Notes). Full-text article PDFs were converted into markdown (text-based formatting) with the Gemini 2.0 Flash model for full-text screening.

#### Screening Evaluation Methodology

We retrospectively evaluated screening performance using a diagnostic test accuracy study design. **Our reference standard was the final article inclusion or exclusion decisions of the original review authors after full-text screening**. “Included” articles represented the final set of articles included in each review, and “excluded” articles represented articles excluded from title, abstract, and full-text screening in each review.

We tested the *otto-SR* screening agent as a standalone reviewer across the full set of citations retrieved in all five SRs (n = 32,357 total citations) (Table 3,4). For comparison, we assessed the screening performance of three commercially available systematic review automation softwares (Elicit, Nested Knowledge, DistillerSR; evaluated January 2026), and postgraduate human researchers (n=4) with past SR experience, against a representative sample of citations from each SR (n=1,767 total citations) (Table 3,4; Supplementary Methods).

#### Human screening calibration

To validate the proficiency of our human reviewers in screening, we conducted a calibration exercise (n=400 citations). Our reviewers’ screening decisions were compared with those from an independent re-screening conducted by the original (SeroTracker) study authors, which was completed for a prior validation study.^[19]^

In that study,^[19]^ the SeroTracker authors re-screened articles that had already been included or excluded as part of their living systematic review. The original SeroTracker screening decisions (prior to re-screening; generated through the initial review process) served as the reference standard for this calibration exercise, while the re-screening provided an external benchmark of expert screening performance. Our team of human reviewers had similar performance to the independent re-screening by the SeroTracker study authors (Our team: 80.2% sensitivity 97.7% specificity vs. re-screening Serotracker team: 81.3% sensitivity, 98.1% specificity) providing confidence that our reviewers had expert-level screening proficiency (Table 2).

**Table 2.** Screening calibration: Performance of human reviewers relative to original study authors.

| Screening Stage | Group | True positives, n | True negatives, n | False positives, n | False negatives, n | Sensitivity | Specificity |
| --- | --- | --- | --- | --- | --- | --- | --- |
| Dual Abstract | SeroTracker Human | 77 | 297 | 12 | 4 | 84.6% | 96.1% |
|  | Team 1* | 75 | 299 | 10 | 16 | 82.4% | 96.8% |
|  | Team 2* | 78 | 296 | 13 | 13 | 85.7% | 95.8% |
| Dual full-text | SeroTracker Human | 74 | 299 | 10 | 17 | 81.3% | 96.8% |
|  | Team 1* | 72 | 310 | 8 | 19 | 79.1% | 97.4% |
|  | Team 2* | 74 | 303 | 6 | 17 | 81.3% | 98.1% |

\*Team 1 and Team 2 represent our panel of human reviewers, who screened records independently and in duplicate. Conflicts were arbitrated by a third reviewer

**Table 3.**
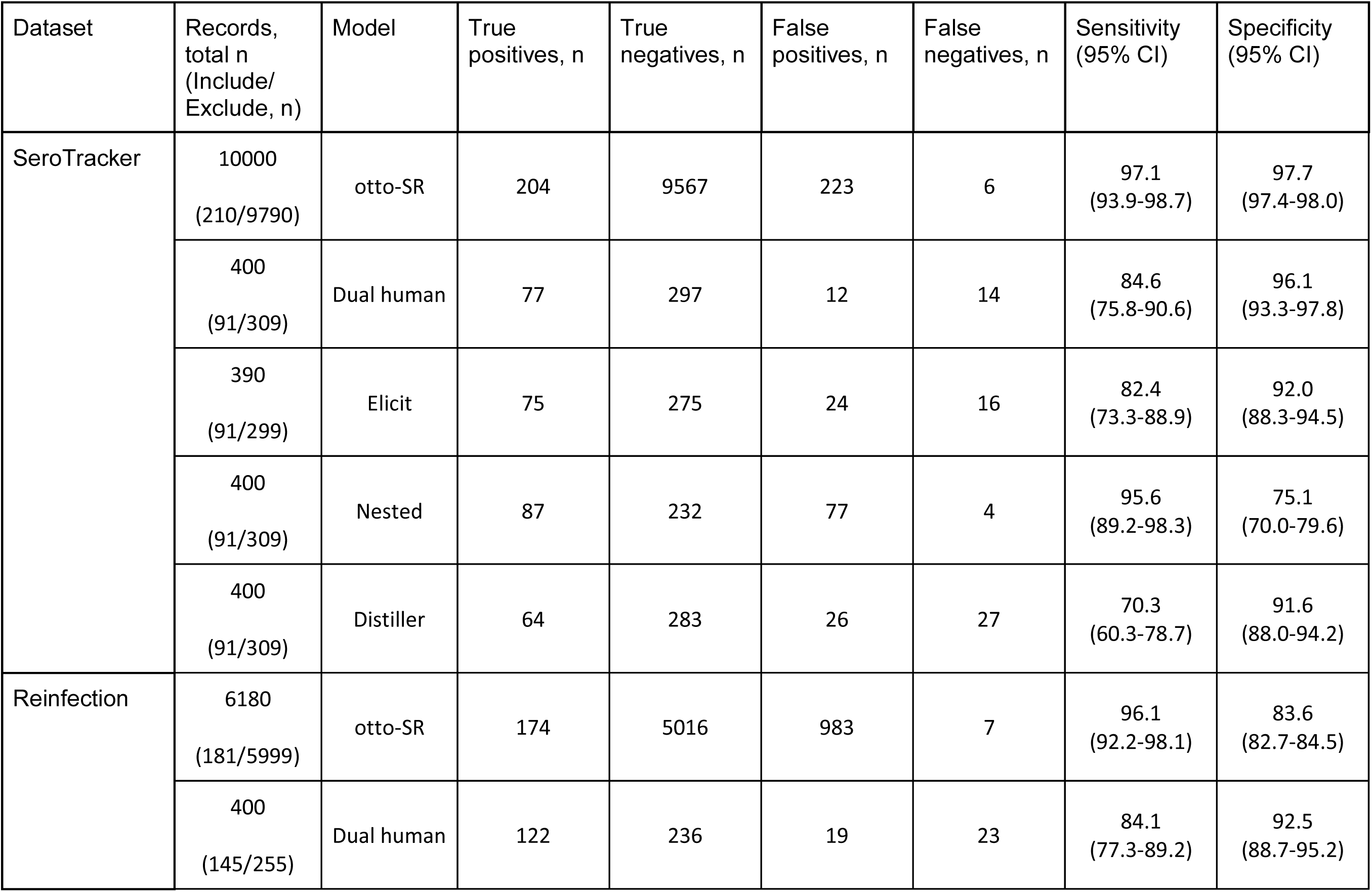
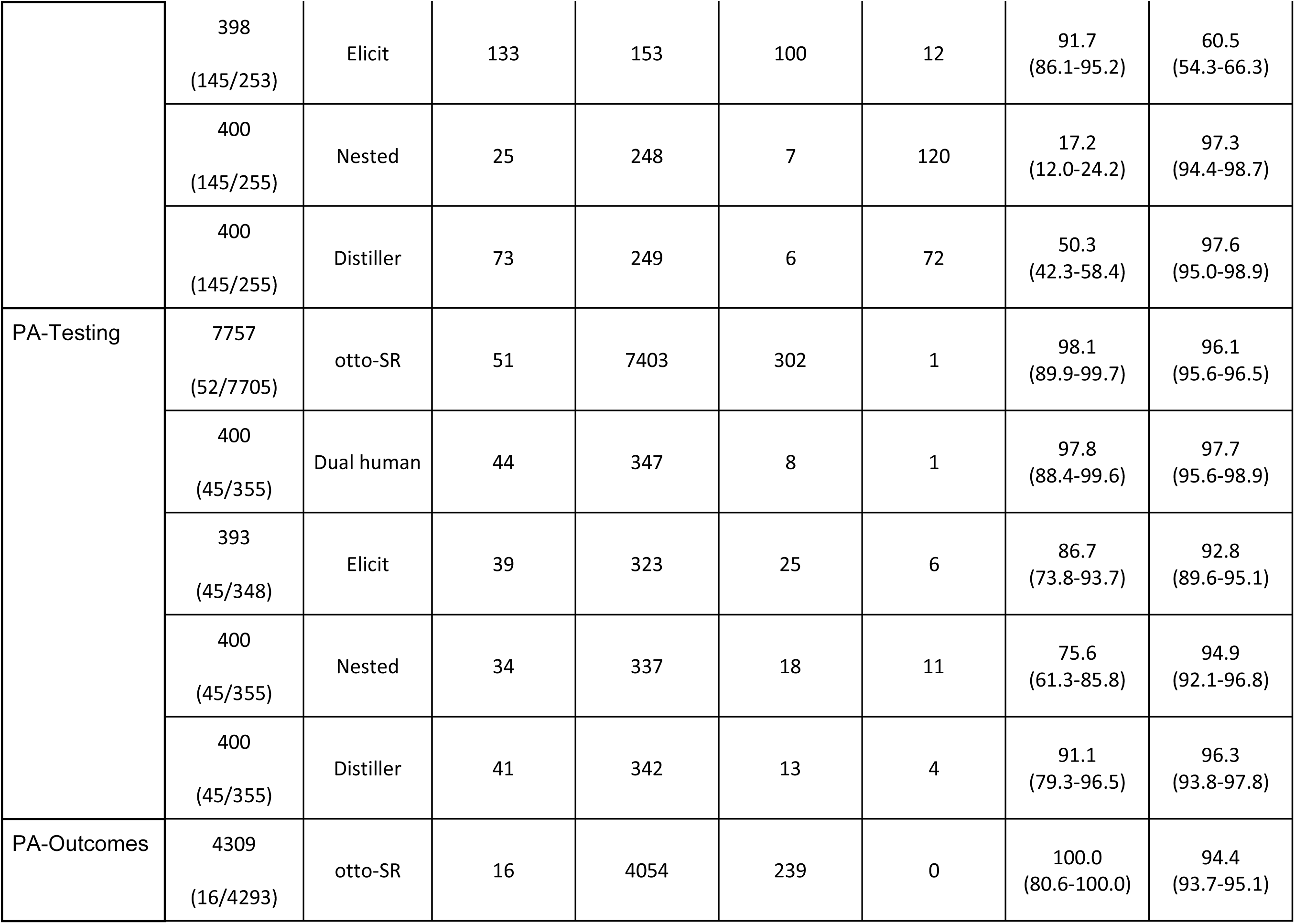
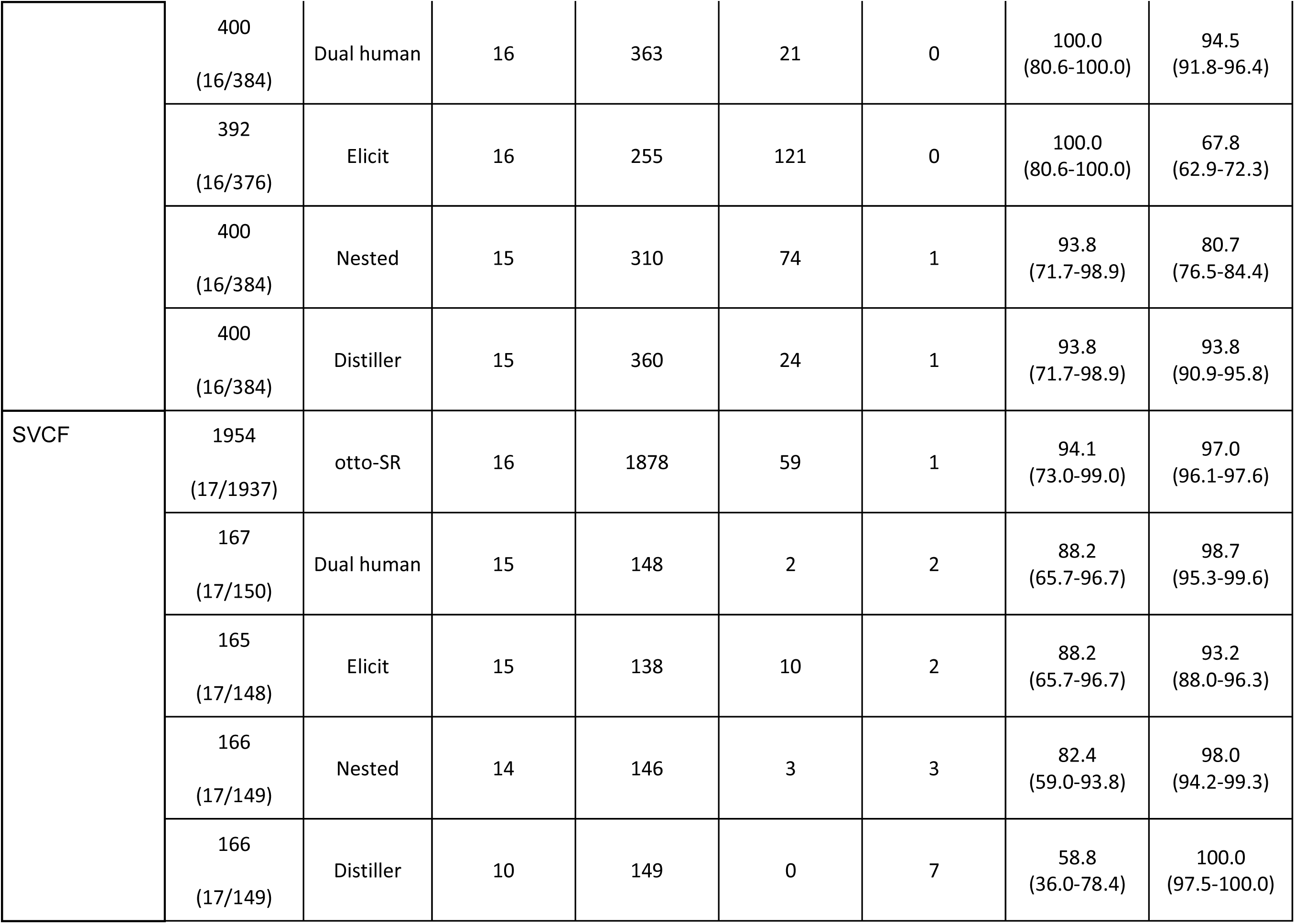
Phase 1: Abstract screening performance of *otto-SR*, dual human reviewers, Elicit, NestedKnowledge, and DistillerSR.

#### Screening Data Analysis

We assessed the performance of the *otto-SR* screening agent, Elicit, Nested Knowledge, DistillerSR, and the external dual human reviewers by measuring accuracy, sensitivity, and specificity. We calculated 95% CIs for weighted (pooled-denominator) sensitivity and specificity using the Wilson method^[20]^ with the binom package in R.

### Phase 2: Evaluation of Systematic Review Data Extraction Performance

We gathered four datasets (SeroTracker,^[14]^ PA-Outcomes,^[16]^ PA-Testing,^[17]^ Sepsis^[21]^) from the *BenchSR* database and three external SRs (CKD,^[22]^ Process,^[23]^ Psyc-meds^[24]^) via convenience sampling that provided publicly accessible raw data extraction information (see Table 5 for details of reviews). Variables for data extraction included descriptive information and outcome details (see Supplementary Notes).

**Table 4.** Phase 1: Full-text screening performance of *otto-SR* and dual human reviewers.

| Dataset | Records,<br>total n<br>(Include/<br>Exclude, n) | Model | True<br>positives, n | True<br>negatives, n | False<br>positives, n | False<br>negatives, n | Sensitivity<br>(95% CI) | Specificity<br>(95% CI) |
| --- | --- | --- | --- | --- | --- | --- | --- | --- |
| Serotracker | 10000<br>(210/9790) | otto-SR | 204 | 9663 | 127 | 6 | 97.1<br>(93.9-98.7) | 98.7<br>(98.5-98.9) |
|  | 400<br>(91/309) | Dual human | 74 | 299 | 10 | 17 | 81.3<br>(72.1-88.0) | 96.8<br>(94.1-98.2) |
|  | 390<br>(91/299) | Elicit | 72 | 284 | 15 | 19 | 79.1<br>(69.7-86.2) | 95.0<br>(91.9-96.9) |
|  | 400<br>(91/309) | Nested | 85 | 266 | 43 | 6 | 93.4<br>(86.4-96.9) | 86.1<br>(81.8-89.5) |
| Reinfection | 6180<br>(181/5999) | otto-SR | 174 | 5439 | 560 | 7 | 96.1<br>(92.2-98.1) | 90.7<br>(89.9-91.4) |
|  | 400<br>(145/255) | Dual human | 64 | 243 | 12 | 81 | 44.1<br>(36.3-52.3) | 95.3<br>(92.0-97.3) |
|  | 398<br>(145/253) | Elicit | 132 | 180 | 73 | 13 | 91.0<br>(85.3-94.7) | 71.1<br>(65.3-76.4) |
|  | 400<br>(145/255) | Nested | 21 | 248 | 7 | 124 | 14.5<br>(9.7-21.1) | 97.3<br>(94.4-98.7) |
| PA-Testing | 7757<br>(52/7705) | otto-SR | 48 | 7608 | 97 | 4 | 92.3<br>(81.8-97.0) | 98.7<br>(98.5-99.0) |
|  | 400<br>(45/355) | Dual human | 36 | 351 | 4 | 9 | 80.0<br>(66.2-89.1) | 98.9<br>(97.1-99.6) |
|  | 393<br>(45/348) | Elicit | 38 | 327 | 21 | 7 | 84.4<br>(71.2-92.3) | 94.0<br>(91.0-96.0) |
|  | 400<br>(45/355) | Nested | 33 | 338 | 17 | 12 | 73.3<br>(59.0-84.0) | 95.2<br>(92.5-97.0) |
| PA-Outcomes | 4309<br>(16/4293) | otto-SR | 16 | 4179 | 114 | 0 | 100.0<br>(80.6-100.0) | 97.3<br>(96.8-97.8) |
|  | 400<br>(16/384) | Dual human | 15 | 376 | 8 | 1 | 93.8<br>(71.7-98.9) | 97.9<br>(95.9-98.9) |
|  | 392<br>(16/376) | Elicit | 16 | 269 | 107 | 0 | 100.0<br>(80.6-100.0) | 71.5<br>(66.8-75.9) |
|  | 400<br>(16/384) | Nested | 15 | 317 | 67 | 1 | 93.8<br>(71.7-98.9) | 82.6<br>(78.4-86.0) |
| SVCF | 1954<br>(17/1937) | otto-SR | 16 | 1902 | 35 | 1 | 94.1<br>(73.0-99.0) | 98.2<br>(97.5-98.7) |
|  | 167<br>(17/150) | Dual human | 13 | 150 | 0 | 4 | 76.5<br>(52.7-90.4) | 100.0<br>(97.5-100.0) |
|  | 165<br>(17/148) | Elicit | 14 | 140 | 8 | 3 | 82.4<br>(59.0-93.8) | 94.6<br>(89.7-97.2) |
|  | 166<br>(17/149) | Nested | 13 | 146 | 3 | 4 | 76.5<br>(52.7-90.4) | 98.0<br>(94.2-99.3) |

Screening decisions following abstract screening then underwent full-text screening.

**Table 5.** Phase 2: Descriptive overview of datasets used in data extraction benchmark.

| Dataset | Clinical Domain (Web of Science) | Population | Intervention/ Exposure | Study Types Included | Total articles<br>[Sample N]* | Total data-points<br>[Sample N]* |
| --- | --- | --- | --- | --- | --- | --- |
| SeroTracker<br>*Test sample | Infectious Diseases | Humans of any age | Prevalence of SARS-CoV-2 antibodies | Cross-sectional, repeated cross sectional, and cohort study designs | 100<br>[30] | 700<br>[210] |
| PA-Testing | Endocrinology & Metabolism | Patients with primary aldosteronism | Confirmatory tests used to diagnose primary aldosteronism | Case-control, diagnostic testing | 51<br>[12] | 867<br>[204] |
| PA-Outcomes | Endocrinology & Metabolism<br>Surgery | Patients with primary aldosteronism | Surgical adrenalectomy and medical treatment with a mineralocorticoid receptor antagonist | Randomized controlled trials, observational studies | 16<br>[10] | 336<br>[210] |
| Sepsis | General & Internal Medicine | Individuals aged $\geq 16$ years with septic shock (sepsis and use of at least one vasopressor) | Treatment with hydrocortisone alone, hydrocortisone-fludrocortisone, placebo or usual care. | Randomized controlled trials | 16<br>[13] | 256<br>[208] |
| CKD | Urology & Nephrology | Empirical literature studying nature or burden of CKD progression | N/A | Retrospective cohort studies, case-control studies, cross-sectional studies | 80<br>[21] | 800<br>[210] |
| Process | Health Care Sciences & Services | Empirical literature studying process mapping in healthcare | N/A | Methodological studies | 104<br>[17] | 1248<br>[204] |
| Psyc-meds | Pharmacology & Pharmacy<br>Psychiatry | Adults with a diagnosis of generalized anxiety disorder | Antidepressant monotherapy | Randomized controlled trials | 28<br>[23] | 252<br>[207] |

\*Sample N refers to the number of studies that underwent data extraction by dual human reviewers and DistillerSR. The sample size was based on a McNemar test sample size approximation.

#### Development of LLM-based Data Extraction Agent

We developed an extraction agent using the OpenAI o3-mini-high model.^[25]^ In all cases, the *otto-SR* extraction agent was prompted with original author-defined variable descriptions. Full-text article PDFs were converted into markdown format with the Gemini 2.0 Flash model.

#### Data Extraction Evaluation Methodology

We retrospectively measured the performance of the *otto-SR* data extraction agent, Elicit, and Nested Knowledge in data extraction across all seven SRs (n=4,459 data points, 495 studies) (Table 5). The performance of DistillerSR and graduate-level human reviewers were assessed on a randomly sampled subset of articles from each review. The sample size was based on a McNemar test sample size approximation (n=1,453 data points, 156 studies) (Supplementary Methods). Graduate human reviewers performed data extraction in duplicate, with arbitration by a third reviewer.

Data extraction accuracy was determined through an LLM-as-a-judge framework to compare AI-or human extracted values (graduate-level reviewers) against the original author extractions (Supplementary Methods). However, given the known variability in dual human data extraction accuracy (reported rates: 65.8-85.5%),^[26–31]^ original author-extracted values were not treated as a definitive gold standard. Instead, we applied a blinded adjudication process to resolve discrepancies between *otto-SR* extraction and the original authors. A panel of three blinded human reviewers compared randomized pairs of responses (*otto-SR* vs. original author) and selected the most accurate value (Supplementary Methods). **We used these final adjudicated judgements to construct a corrected reference standard** for all performance evaluations. This adjudication framework was adapted from established protocols in radiology, ophthalmology, and clinical trials.^[32–35]^

We additionally performed a post-hoc review of incorrect AI extraction to classify errors as either ‘parsing errors’ (i.e., errors with converting PDF files to markdown formatting), ‘inaccessible’ (i.e., data accessible only through author correspondence or supplementary material), or ‘true errors’ (i.e., cases where the original author values were incorrectly extracted).

#### Data Extraction Analysis

We assessed the performance of the *otto-SR* data extraction agent, Elicit, Nested Knowledge, DistillerSR, and external human reviewers by measuring the total accuracy at a variable level per study. If the human adjudicator classification was “inaccessible”, the data point was removed from analysis. We calculated 95% CIs for weighted (pooled-denominator) accuracy using the Wilson method^[20]^ with the binom package in R.

### Phase 3: Evaluation of Systematic Review Risk of Bias Assessment Performance

We gathered 12 SR datasets via convenience sampling that reported publicly accessible risk of bias assessments using Cochrane RoB2^[36]^ for RCTs (n=4 SRs, 214 studies),^[37–40]^ Newcastle-Ottawa-Scale^[41]^ for cohort studies (n=4 SRs, 75 studies),^[42–45]^ and QUADAS2^[46]^ for diagnostic test accuracy studies (n=4 SRs, 56 studies).^[47–50]^

#### Development of an LLM-based Risk of Bias Agent

We developed a risk of bias (ROB) agent using the OpenAI GPT-5 model. In all cases, the *otto-SR* ROB agent was prompted with the full critical appraisal tool instructions as published by the creators (ROB2, NOS, or QUADAS2), and review dataset metadata required for appraisal (e.g., outcomes assessed, objectives).

#### Risk of Bias Evaluation Methodology

We compared the performance of the otto-SR RoB agent with original review author appraisals for each validated tool (e.g., ROB2). As risk of bias is inherently subjective and prone to interpretive variability, we applied a blinded adjudication process involving independent reviewers to resolve disagreements between otto-SR and author assessments. **The resulting adjudicated judgements were used to construct a corrected reference standard.** Both *otto-SR* and original review author decisions were evaluated against this corrected reference standard.

#### Risk of Bias Analysis

We assessed the performance of the *otto-SR* RoB agent by measuring the total accuracy of risk of bias assessments (number of concordant appraisal assessments for each domain/question). We report confusion matrices (per domain; overall RoB) and inter-rater reliability metrics, including Gwet’s AC2, which was selected for its robustness to category prevalence and class imbalance (Supplementary Methods). We calculated 95% CIs for weighted (pooled-denominator) accuracy using the Wilson method^[20]^ with the binom package in R.

### Phase 4: Evaluation of otto-SR for Reproducing and Updating Cochrane Reviews

To evaluate the reliability and generalizability of our automated SR workflow, we conducted a focused reproducibility assessment using one complete issue of the Cochrane Database of Systematic Reviews. Our aim was to reproduce each review’s workflow end-to-end, from literature search to meta-analysis, while using the *otto-SR* pipeline for screening, extraction, and RoB assessments.

We followed Cochrane methodology with one exception. Cochrane reviews typically include studies even if they do not report data for the review’s primary outcome. This method allows for reporting of all outcomes for potential comparisons (e.g., all intervention and outcome combinations). In contrast, we focused on reproducing each review’s predefined primary outcome. This constraint provided a clearer distinction for study eligibility.

We randomly selected the April 2024 issue of the Cochrane Database (Table 8, Supplementary Methods).^[51–64]^ Of the 14 reviews in this issue, one review was excluded due to a lack of publicly available data, and a second review was excluded due to the absence of a reproducible search strategy.

#### Cochrane Database Searches

The original search strategy of each Cochrane review was reproduced using the exact terms and filters described in the review methods. Searches were limited to institutionally accessible databases. In cases where databases lacked precise date filtering (e.g., supporting month but not day-level granularity), we applied post-hoc filtering to approximate the original search window (Supplementary Data 2).

After each search, we cross-referenced our list of articles with those included in the original Cochrane reviews. Articles that were not retrievable using the original database search strategies were excluded from downstream screening, data extraction and analysis.

#### Cochrane Screening

All retrieved citations underwent abstract and full-text screening with the *otto-SR* screening agent, prompted with the original objectives, and inclusion and exclusion criteria from each Cochrane review (Supplementary Notes).

We retrieved additional abstract or DOI information from incomplete citation records through OpenAlex.^[65]^ Citations without a retrievable abstract or DOI/trial identifier were excluded.

Screening decisions were compared against the original Cochrane author decisions to calculate true positives, true negatives, false negatives, and false positives.

#### Cochrane Data Extraction

All studies included after full-text screening underwent data extraction using the *otto-SR* extraction agent which focused on the primary outcome defined in each Cochrane review. The extraction prompts used the original author-defined variable names and extraction logic (Supplementary Notes).

Data extraction also served as a final screening filter. Studies were programmatically excluded if they returned unavailable or unreportable values for the primary outcome (e.g., “na”), were identified as duplicates, or involved only ineligible intervention-comparator pairs (e.g., Drug A vs. Drug B when only Drug A vs. placebo was eligible).

#### Cochrane Statistical Analysis

All meta-analyses were conducted using the metafor package in R (Code and Dataset Availability). We matched the original authors’ specified meta-analytic model (e.g., random-effects, fixed-effect), effect size metric (risk ratio, odds ratio, rate ratio, mean difference, standardized mean difference), and continuity correction approach, where reported.

We conducted four meta-analytical comparisons: (1) ***Cochrane*** – meta-analysis using the original author-extracted data; (2) ***Matched*** – *otto-SR* results filtered to match the studies included by the original Cochrane authors; (3) ***Expanded*** – all eligible studies included by *otto-SR* under the original search date cut-off including those not in the original Cochrane review; (4) ***Updated*** – all eligible studies included by *otto-SR* using an updated search date extending to May 8 2025. We also derived ‘corrected’ values, for the ‘matched,’ ‘expanded,’ and ‘updated’ comparisons that served as the reference ground truth for analytical comparison.

#### Cochrane Data Correction and Comparison

Given potential data extraction errors by original Cochrane authors and *otto-SR*, we derived corrected values for each analytical comparison through dual human review. This included removal of false positive articles and inclusion of false negative articles, as well as correction of extracted data points. **All original Cochrane data, *otto-SR* extracted data, and corrected data (including descriptions of why data were corrected) are provided in Supplementary Data 1.** Details on our data correction process are provided in the Supplementary Methods.

#### Cochrane Risk of Bias assessment

We applied the otto-SR RoB agent to the articles included in the ‘matched’ analysis, restricting inclusion to reviews using ROB2, Newcastle Ottawa, or QUADAS2 tools. Details on our risk of bias assessment and correction process is provided in the Supplementary Methods.

## Results

### Phase 1: Evaluation of Screening Performance

At the abstract screening stage, the *otto-SR* screening agent achieved the highest sensitivity (weighted sensitivity 96.6% [total range, 94.1-100.0%]) (Fig 2, Table 3) followed by Elicit (88.5% [76.9-100%] sensitivity), dual human reviewers (87.3% [84.1-100%] sensitivity), DistillerSR (61.4% [50.3-93.8%] sensitivity), and Nested Knowledge (55.7% [17.2-95.6%] sensitivity). Dual human reviewers achieved the highest specificity in abstract screening (95.7% [92.5-98.7%] specificity), followed by DistillerSR (95.3% [91.6-100%] specificity), the *otto-SR* screening agent (93.9% [83.6-97.7%] specificity), Nested Knowledge (87.7% [75.0-98.0%] specificity), and Elicit (84.2% [65.7-95.9%] specificity).

**Figure 2.**
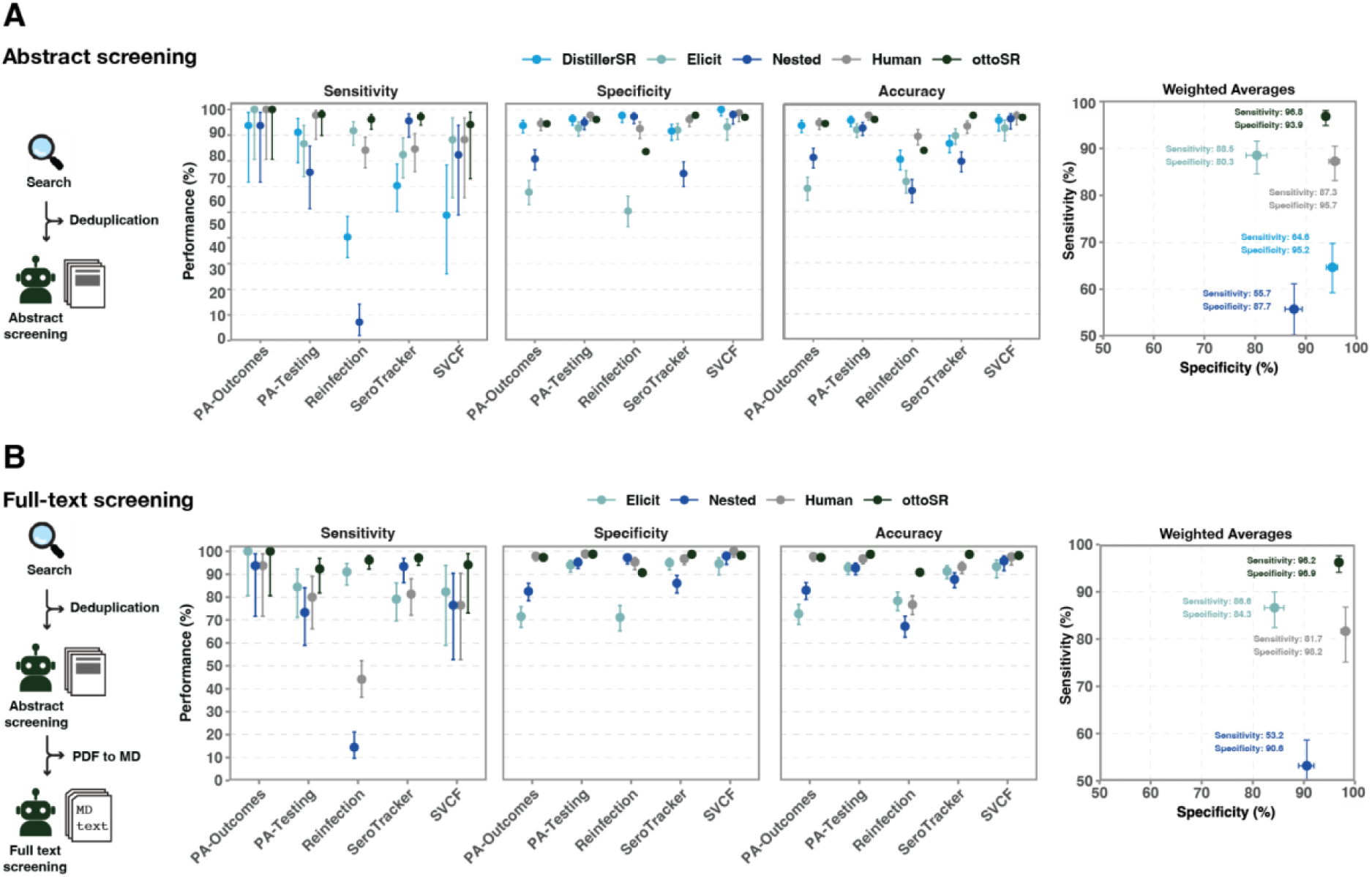
*otto-SR* screening agent performance on systematic review screening. **A.** diagram of *otto-SR* **abstract screening** agent (left), sensitivity, specificity of *otto-SR* screening agent, dual human reviewers, Elicit, Nested Knowledge, and DistillerSR for abstract screening evaluated across five reviews (middle). Weighted average for sensitivity and specificity across comparator groups (five reviews) (right). Error bars indicate 95% confidence intervals. **B.** diagram of *otto-SR* **full-text screening** agent (left), sensitivity, specificity, of *otto-SR* screening agent, dual human reviewers, Elicit, and Nested Knowledge for full-text screening evaluated across five reviews (middle). Weighted average for sensitivity and specificity across otto-SR, Elicit, Nested Knowledge (five reviews) and dual human (four reviews) (right). Error bars indicate 95% confidence intervals. Abbreviations: Markdown file (MD).

After full-text screening, the *otto-SR* screening agent maintained the highest sensitivity (96.2% [92.3-100%] sensitivity), followed by Elicit (86.6% [79.12-100%] sensitivity), while human reviewers had a marked drop in sensitivity (63.3% [44.1-93.8%] sensitivity) (Fig 2, Table 4). This decline in human sensitivity was largely driven by poor performance on screening the “Reinfection” review (44.1% sensitivity, 95.3% specificity), likely due to complex inclusion criteria involving test-negative study designs, multiple interventions, and multiple time-specific outcomes. After removing this outlier review, human reviewers achieved a weighted sensitivity of 81.7% [76.4%-93.8%]. Nested Knowledge had the lowest full-text screening sensitivity of all four groups tested (53.2% [14.5-93.8%] sensitivity). Specificity remained high for both the *otto-SR* screening agent (96.9% [90.7-98.7%] specificity) and dual human reviewers (98.1% [96.7-100.0%] specificity). Elicit had the lowest specificity among all four groups tested (86.6% [82.6-98.0%] specificity).

### Phase 2: Evaluation of Data Extraction Performance

Across all seven reviews, the *otto-SR* extraction agent achieved an average weighted accuracy of 93.1% (91.1.4-97.0%), outperforming dual human reviewers at 79.7% (69.1-91.0%), Elicit at 76.0% (60.7-85.8%), Nested Knowledge at 71.2% (39.1-83.8%), and DistillerSR at 46.1% (29.9-63.2%) (Fig 3a, Table 6). Notably, our human reviewers performed comparably to prior estimates of human data extraction accuracy (Fig 3b). When *otto-SR* extracted different values to the original authors, the blinded human reviewers sided with the *otto-SR* data extraction agent in 69.3% of cases (Fig 3c). In contrast, for discrepancies between original authors and our two human extractors, the blinded reviewer panel sided with the dual human extractors in 28.1% of cases.

**Figure 3.**
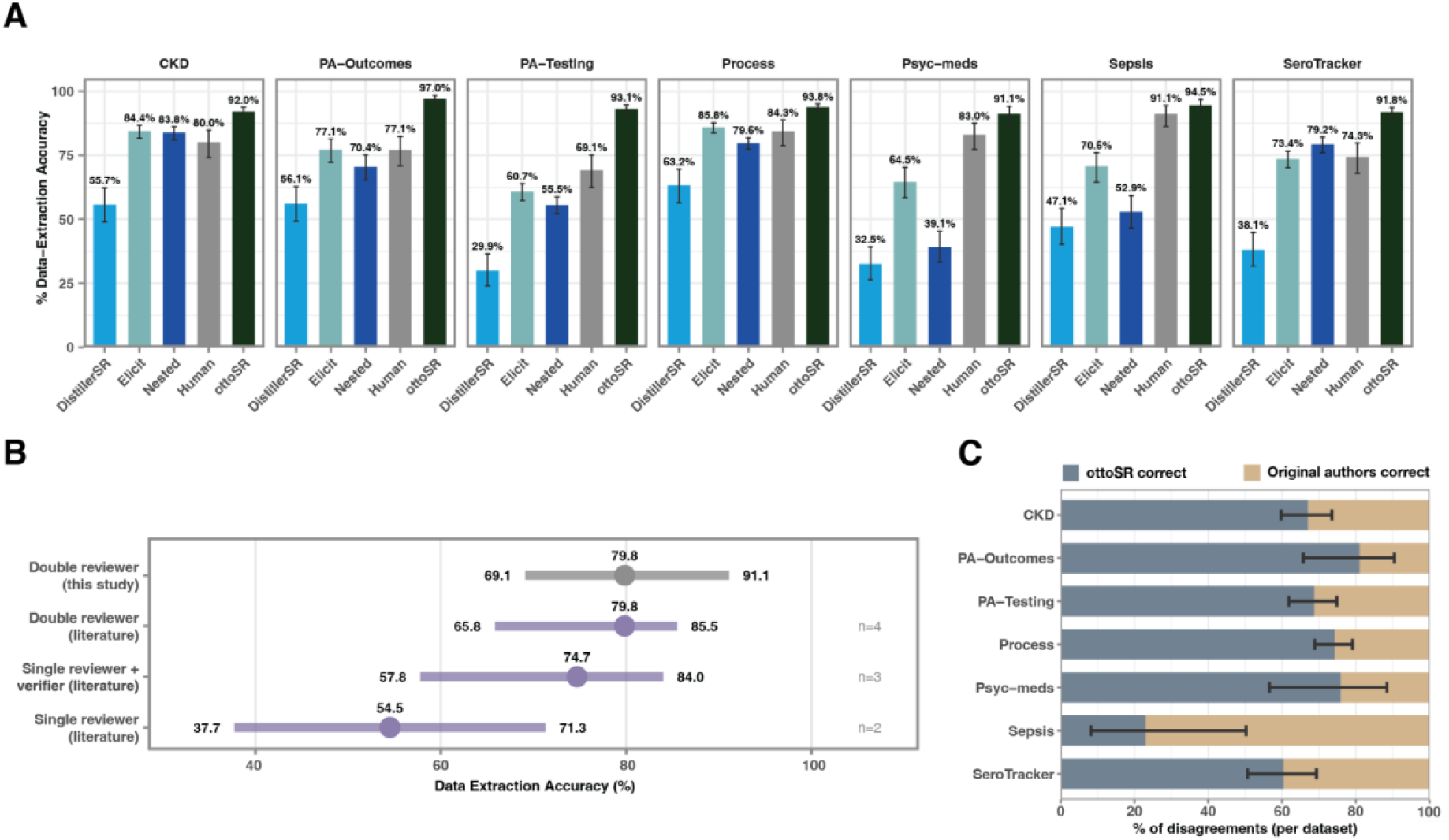
*otto-SR* extraction agent performance on systematic review data extraction. **A.** bar graph displaying data extraction accuracy of the *otto-SR* extraction agent (green) (4459 data points), Elicit (teal) (4459 data points), Nested Knowledge (dark blue) (4459 data points), DistillerSR (light blue) (1,453 data points), and dual human reviewers (grey) (1,453 data points) across 7 different systematic reviews. Error bars represent 95% confidence intervals. **B.** dot plot depicting literature-derived human reviewer performance comparison (purple) against human reviewers in this study (grey). Dots represent mean value and upper and lower bars represent range. **C.** bar graph depicting dual human adjudicator decisions for values marked as incongruent between original review and *otto-SR* extraction agent and dual human. Blue represents the newly conducted review being correct, while tan represents the original study authors being correct. Error bars represent 95% confidence intervals.

**Table 6.** Phase 2: Accuracy of otto-SR, dual human reviewers, and Elicit in data extraction.

| Dataset | Model | Corrected Numerator | Corrected Denominator | Accuracy |
| --- | --- | --- | --- | --- |
| SeroTracker | Human | 156 | 210 | 74.3 |
|  | Elicit | 509 | 693 | 73.4 |
|  | Otto | 636 | 693 | 91.8 |
|  | Nested | 549 | 693 | 79.2 |
|  | Distiller | 80 | 210 | 38.1 |
| PA-Outcomes | Human | 158 | 205 | 77.1 |
|  | Elicit | 253 | 328 | 77.1 |
|  | Otto | 318 | 328 | 97.0 |
|  | Nested | 231 | 328 | 70.4 |
|  | Distiller | 115 | 205 | 56.1 |
| PA-Testing | Human | 141 | 204 | 69.1 |
|  | Elicit | 526 | 867 | 60.7 |
|  | Otto | 807 | 867 | 93.1 |
|  | Nested | 481 | 867 | 55.5 |
|  | Distiller | 61 | 204 | 29.9 |
| Sepsis | Human | 174 | 191 | 91.1 |
|  | Elicit | 168 | 238 | 70.6 |
|  | Otto | 225 | 238 | 94.5 |
|  | Nested | 126 | 238 | 52.9 |
|  | Distiller | 90 | 191 | 47.1 |
| Process | Human | 172 | 204 | 84.3 |
|  | Elicit | 1071 | 1248 | 85.8 |
|  | Otto | 1170 | 1248 | 93.8 |
|  | Nested | 994 | 1248 | 79.6 |
|  | Distiller | 129 | 204 | 63.2 |
| CKD | Human | 168 | 210 | 80.0 |
|  | Elicit | 675 | 800 | 84.4 |
|  | Otto | 736 | 800 | 92.0 |
|  | Nested | 670 | 800 | 83.8 |
|  | Distiller | 117 | 210 | 55.7 |
| Psyc-meds | Human | 171 | 206 | 83.0 |
|  | Elicit | 160 | 248 | 64.5 |
|  | Otto | 226 | 248 | 91.1 |
|  | Nested | 97 | 248 | 39.1 |
|  | Distiller | 67 | 206 | 32.5 |

In the 6.9% of cases where the *otto-SR* extraction agent was incorrect, post-hoc analysis revealed that 0.83% (37/4459) of data points were inaccessible to the model (supplementary files or data obtained through data request), 0.67% (30/4459) resulted from parsing errors, and 0.49% (22/4459) were cases where neither the *otto-SR* data extraction agent nor original author extraction was correct (Supplementary Fig 1).

### Phase 3: Evaluation of Risk of Bias Assessment Performance

Following blinded adjudication, both the *otto-SR* RoB agent and the original review author assessments were evaluated against a corrected reference standard. Against this reference, *otto-SR* achieved an average weighted accuracy of 96.6% (95% CI: 90.0–97.9) for RoB 2, 94.9% (93.9–97.8) for Newcastle–Ottawa Scale (NOS) cohort assessments, and 78.3% (71.4–84.3) for QUADAS-2 assessments (Figure 4; Table 7, Supplementary Fig. 2). Using the same corrected reference standard, the original review author assessments achieved weighted accuracies of 93.6% (87.1-95.6) for RoB 2, 91.4% (88.9-97.6) for NOS, and 78.3% (53.6-88.3) for QUADAS-2. When *otto-SR* generated different appraisal decisions to the original authors, the blinded human reviewers sided with the *otto-SR* RoB agent in 2.6% of cases (Fig 4). Of note, *otto-SR* had no major discrepancies (i.e., low vs. high ratings) in overall RoB2 domain assessments (Supplementary Fig 3). *otto-SR* also had high interrater reliability, with an average Gwet AC2 of 0.98, 0.95, and 0.74 for ROB2, Newcastle, and QUADAS2 tools, respectively, which was comparable to the agreement between the original review authors (Table 7).

**Figure 4.**
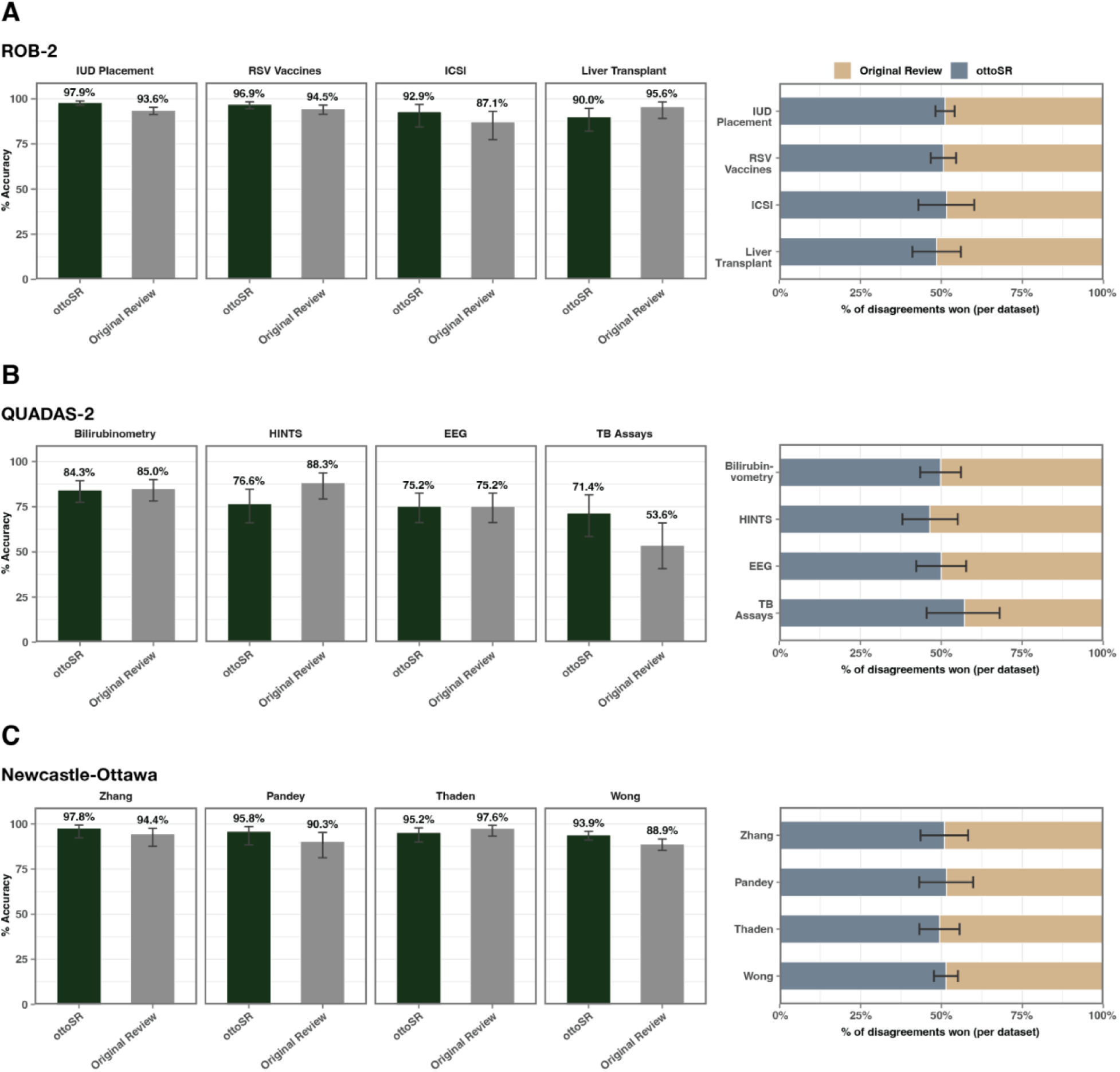
*otto-SR RoB agent* performance on systematic review risk of bias. **A.** Left: bar graph displaying per domain risk of bias accuracy of the *otto-SR* extraction agent (green) (1060 data points) and original review decisions, relative to corrected reference standard across four different systematic reviews with ROB2 risk of bias assessments. Error bars represent 95% confidence intervals. Right: bar graph depicting human adjudicator decisions for values marked as incongruent between original review and *otto-SR RoB agent*. Blue bars indicate instances where the otto-SR risk of bias assessment was adjudicated as correct, while tan bars indicate instances where the original review risk of bias assessment was adjudicated as correct. Error bars represent 95% confidence intervals. **B.** Left: bar graph displaying per domain risk of bias accuracy of the *otto-SR* extraction agent (green) (378 data points) and original review decisions, relative to corrected reference standard across four different systematic reviews with QUADAS2 risk of bias assessments. Error bars represent 95% confidence intervals. Right: bar graph depicting human adjudicator decisions for values marked as incongruent between original review and *otto-SR RoB agent*. Blue bars indicate instances where the otto-SR risk of bias assessment was adjudicated as correct, while tan bars indicate instances where the original review risk of bias assessment was adjudicated as correct. Error bars represent 95% confidence intervals. **C.** Left: bar graph displaying per domain risk of bias accuracy of the *otto-SR* extraction agent (green) (666 data points) and original review decisions, relative to corrected reference standard across four different systematic reviews with Newcastle-Ottawa risk of bias assessments. Error bars represent 95% confidence intervals. Right: bar graph depicting human adjudicator decisions for values marked as incongruent between original review and *otto-SR RoB agent*. Blue bars indicate instances where the otto-SR risk of bias assessment was adjudicated as correct, while tan bars indicate instances where the original review risk of bias assessment was adjudicated as correct. Error bars represent 95% confidence intervals.

**Table 7.** Phase 3: Descriptive overview and risk of bias performance of otto-SR compared to corrected ground truth.

| Dataset | RoB tool | Comparison | Corrected Numerator | Corrected Denominator | Cohen's Kappa | Gwet AC2 |
| --- | --- | --- | --- | --- | --- | --- |
| RSV Vaccines | ROB2 | otto-SR - corrected | 315 | 325 | 0.89 | 0.99 |
|  |  | Original reviewer - corrected | 307 | 325 | 0.76 | 0.98 |
| Liver Transplant | ROB2 | otto-SR - corrected | 81 | 90 | 0.39 | 0.95 |
|  |  | Original reviewer - corrected | 86 | 90 | 0.77 | 0.99 |
| Misoprosotol IUD | ROB2 | otto-SR - corrected | 563 | 575 | 0.86 | 0.99 |
|  |  | Original reviewer - corrected | 538 | 575 | 0.25 | 0.98 |
| Oocyte activation | ROB2 | otto-SR - corrected | 65 | 70 | 0.79 | 0.97 |
|  |  | Original reviewer - corrected | 61 | 70 | 0.56 | 0.94 |
| COVID Diabetes | Newcastle | otto-SR - corrected | 88 | 90 | 0.74 | 0.98 |
|  |  | Original reviewer - corrected | 85 | 90 | -0.03 | 0.94 |
| Sedentary CVD | Newcastle | otto-SR - corrected | 69 | 72 | 0.65 | 0.95 |
|  |  | Original reviewer - corrected | 65 | 72 | 0.18 | 0.89 |
| FUBC Mortality | Newcastle | otto-SR - corrected | 120 | 126 | 0.84 | 0.93 |
|  |  | Original reviewer - corrected | 123 | 126 | 0.92 | 0.97 |
| Smoking surgery complications | Newcastle | otto-SR - corrected | 355 | 378 | 0.65 | 0.93 |
|  |  | Original reviewer - corrected | 336 | 378 | 0.47 | 0.86 |
| Amplitude-integrated EEG | QUADAS2 | otto-SR - corrected | 79 | 105 | 0.55 | 0.66 |
|  |  | Original reviewer - corrected | 79 | 105 | 0.48 | 0.73 |
| HINTS | QUADAS2 | otto-SR - corrected | 59 | 77 | 0.38 | 0.78 |
|  |  | Original reviewer - corrected | 68 | 77 | 0.52 | 0.91 |
| Truenat MTB assay | QUADAS2 | otto-SR - corrected | 40 | 56 | 0.51 | 0.64 |
|  |  | Original reviewer - corrected | 30 | 56 | 0.28 | 0.7 |
| Transcutaneous bilirubinometry | QUADAS2 | otto-SR - corrected | 118 | 140 | 0.57 | 0.87 |
|  |  | Original reviewer - corrected | 119 | 140 | 0.14 | 0.87 |

**Table 8.**
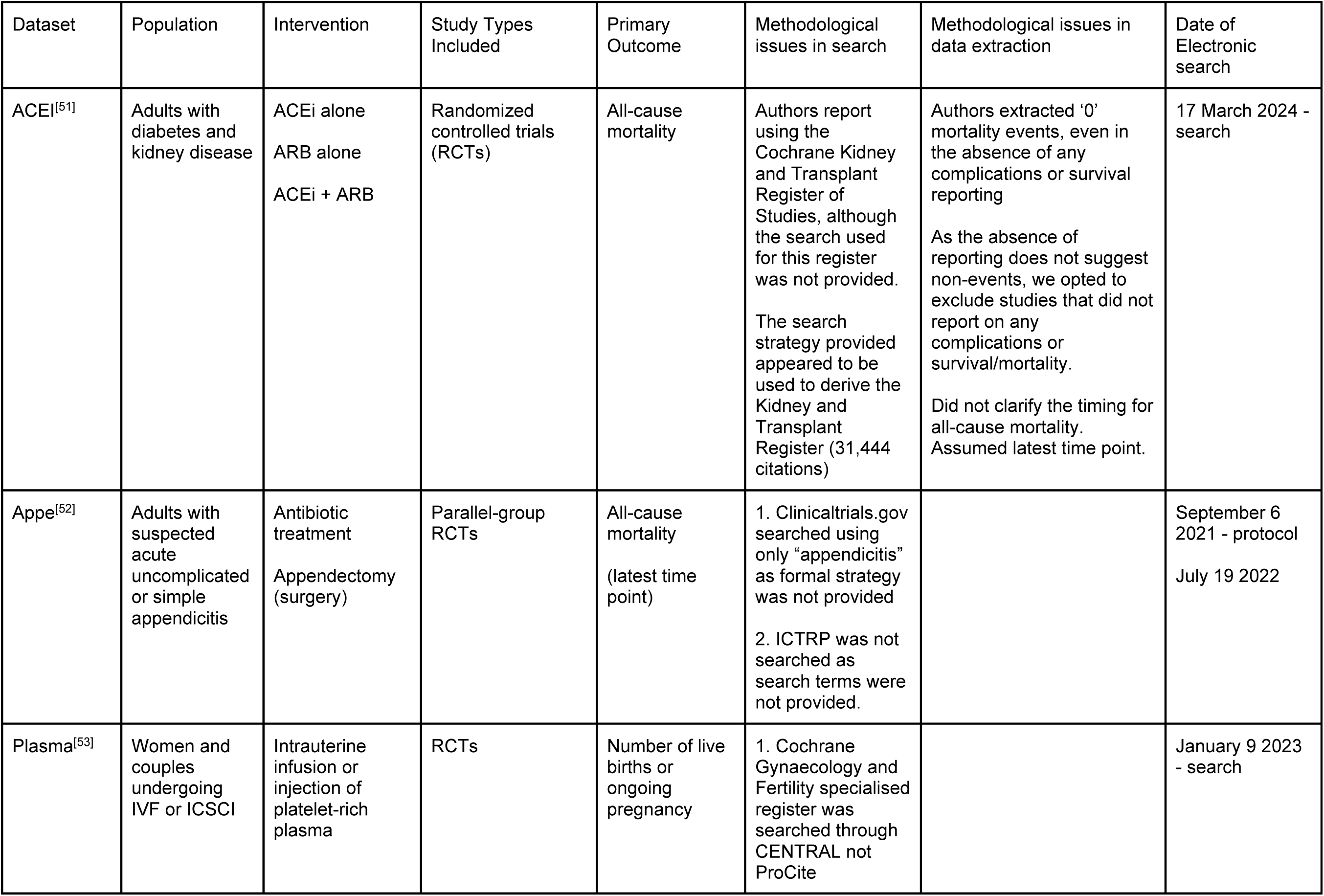
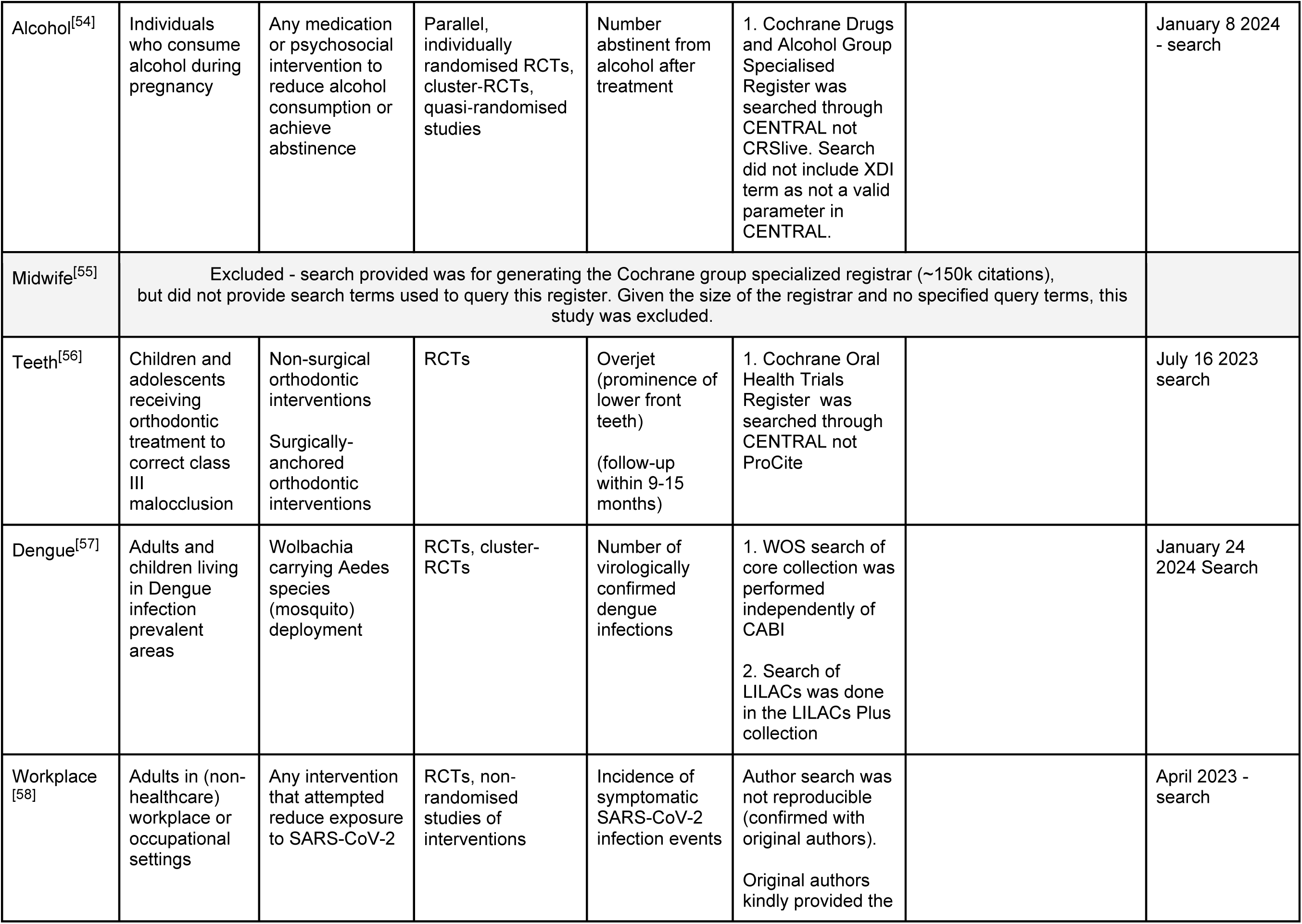
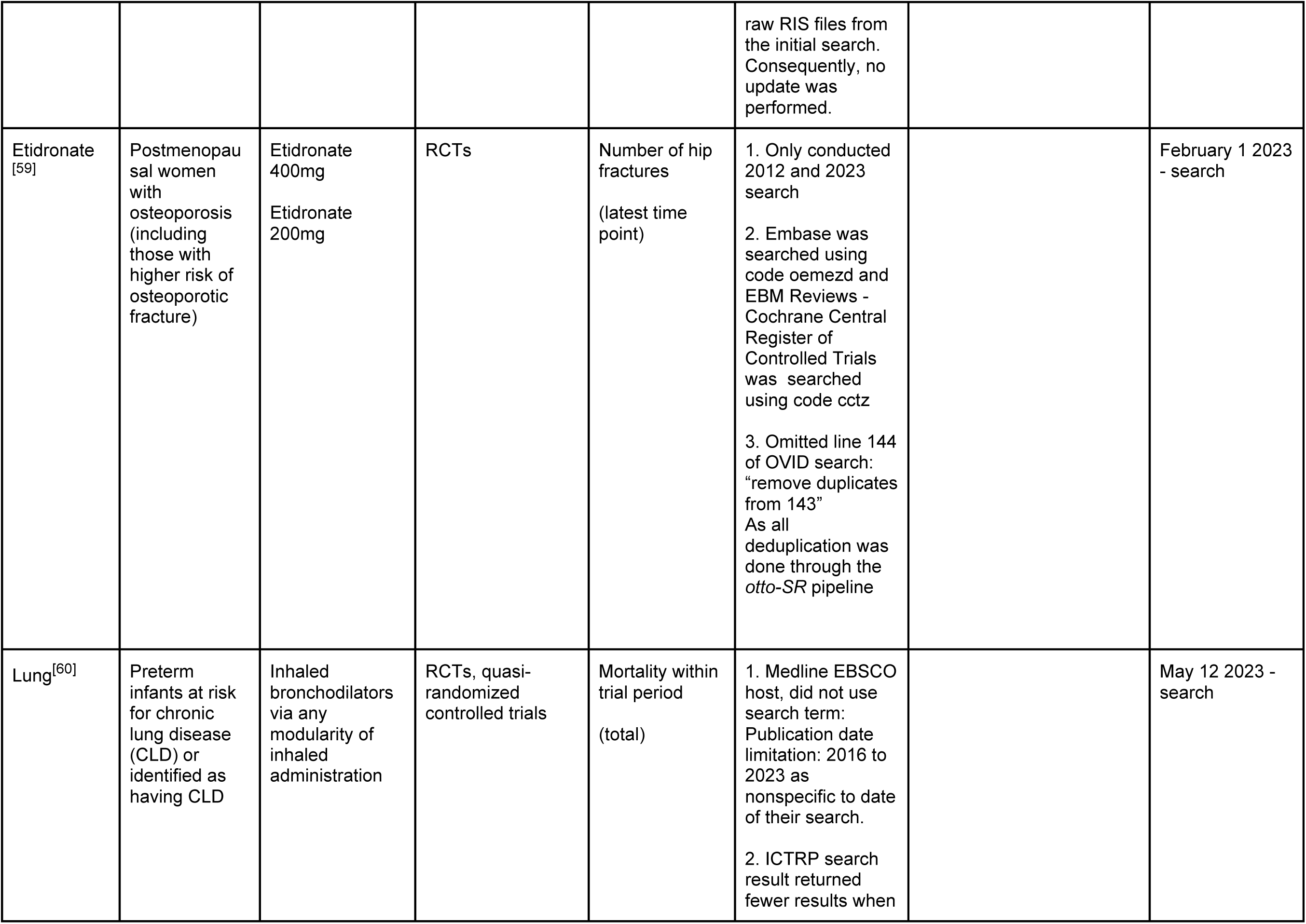
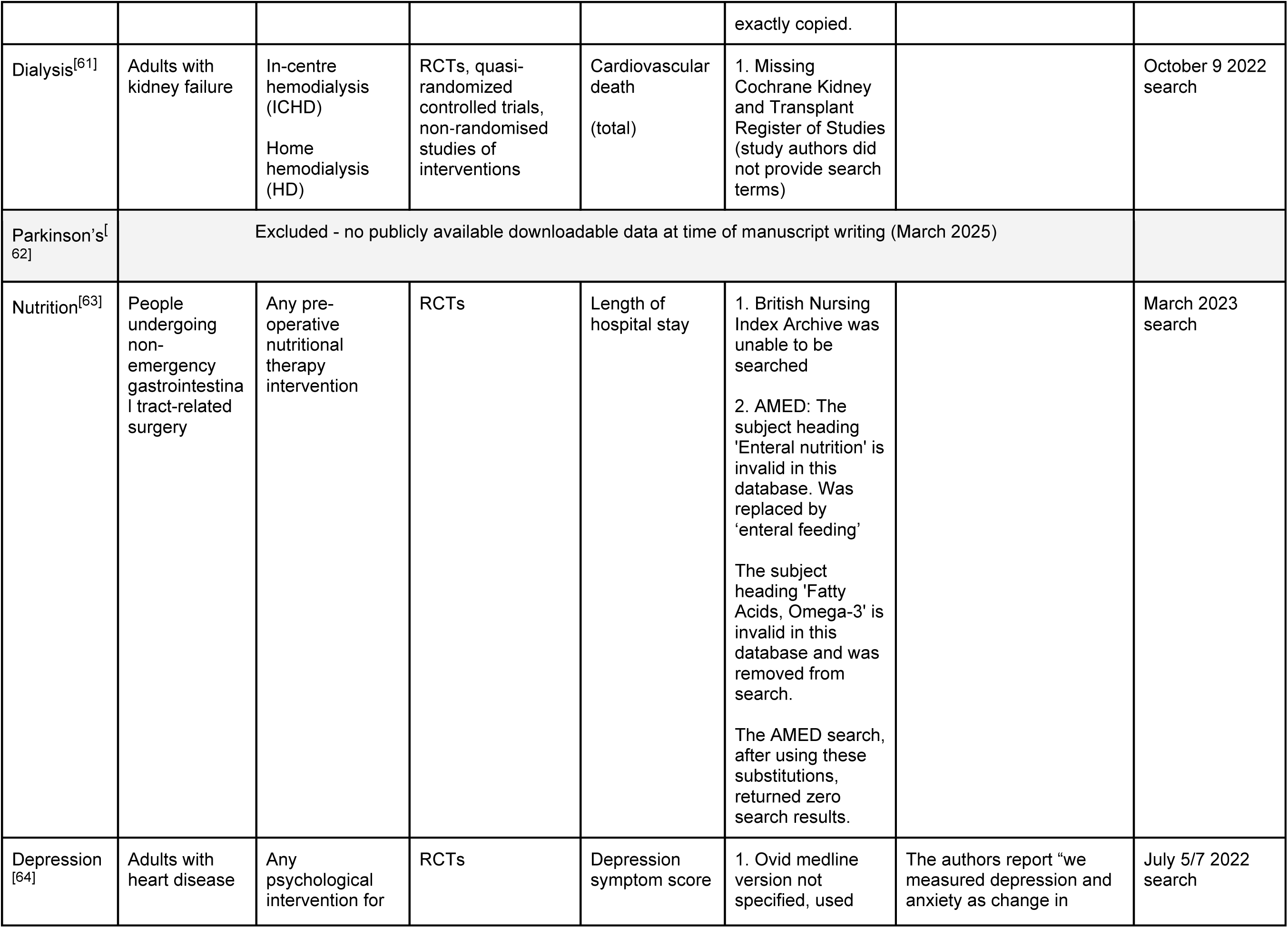
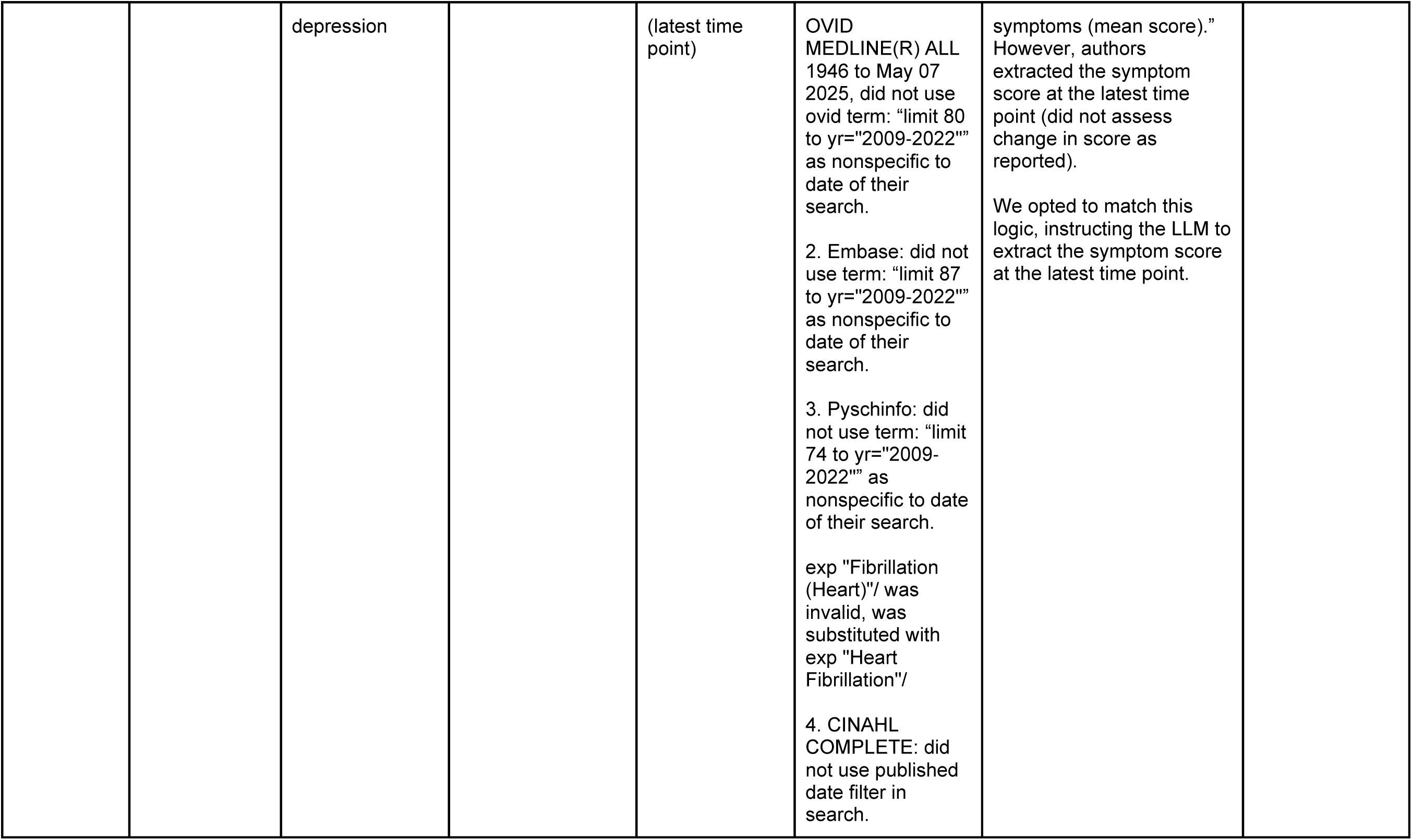
Phase 4: Descriptive overview of reviews included in the Cochrane April 2024 issue.

### Phase 4: Evaluation of otto-SR for Reproducing and Updating Cochrane Reviews

We combined the *otto-SR* screening and extraction agents into a single agentic workflow (Fig 5a) and used it to reproduce and update the primary analysis of 12 reviews from the April 2024 issue of the Cochrane Database of Systematic Reviews (Table 8). We reproduced the reported search strategies, updating searches to May 8, 2025, and identified 146,276 citations.

**Fig 5.**
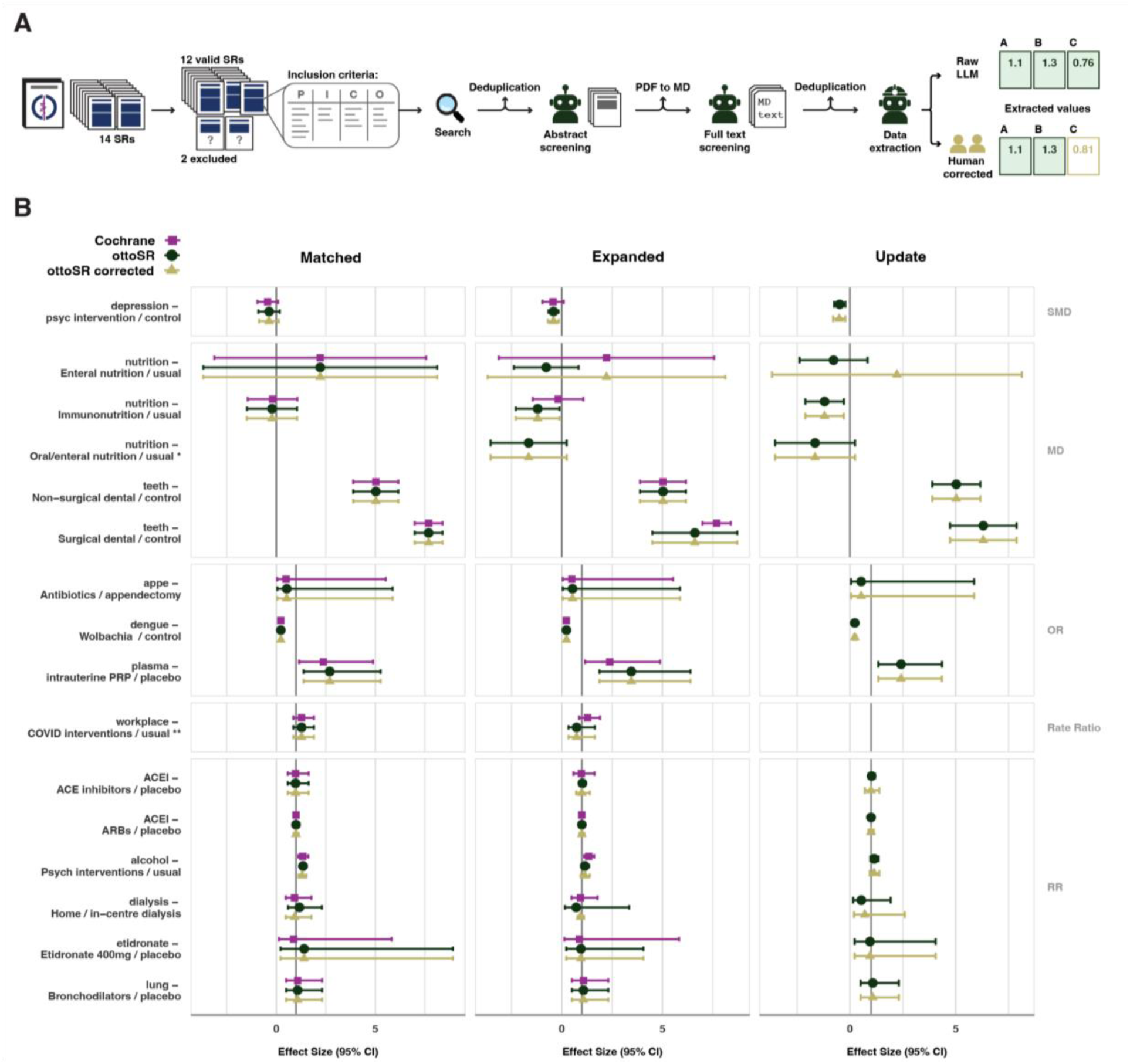
Evaluating *otto-SR* for automation of systematic reviews. A. Infographic depicting use of *otto-SR* for SR automation in a complete edition of the Cochrane Database of Systematic Reviews (n = 12 SRs, n=64 studies). B. Forest plots depicting differences between *otto-SR* (green), original Cochrane study authors (purple), and corrected standard (gold). The ‘corrected’ estimates were derived through dual human review of data extraction values to establish a ground truth. Each row is representative of meta-analyzed estimates derived in a systematic review. Error bars represent 95% confidence interval, MD = Mean Difference, OR = Odds Ratio, RR = Risk Ratio, SMD = Standardized Mean Difference. The **matched** comparison (left) shows estimates derived from articles only included in the original Cochrane reviews. The **expanded** comparison (middle) displays estimates derived from additional articles identified by *otto-SR* falling within the original search dates. The **update** plot (right) displays estimates derived from all articles found by *otto-SR* in a May 8 2025 search. * *otto-SR* discovered a new treatment group, mixed oral / enteral nutrition, which was not found in the original Cochrane review, consequently no matched analysis was conducted. ** workplace citations were provided by original study authors due to challenges with the electronic search, consequently no updated search was performed.

The *otto-SR* screening agent correctly identified all included studies (n=64, 100% sensitivity) across the 12 Cochrane reviews. *otto-SR* extraction results with missing primary outcome values, duplicate studies, or missing intervention-comparator groups were programmatically excluded (Methods). After this process, *otto-SR* incorrectly excluded a median of 0 articles (IQR 0 to 0.25) (Table 9). Incorrect exclusions were due to LLM-inaccessible supplementary data (n=2) or a failure to extract reported outcome values when present (n=2).

**Table 9.**
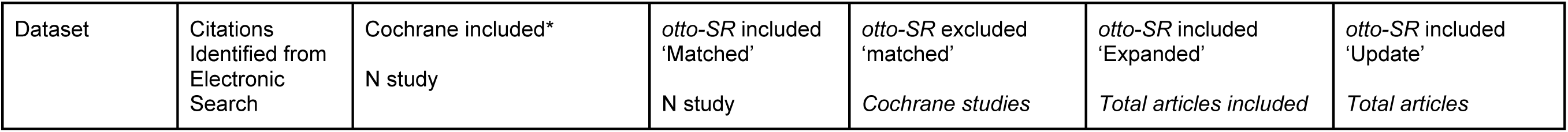
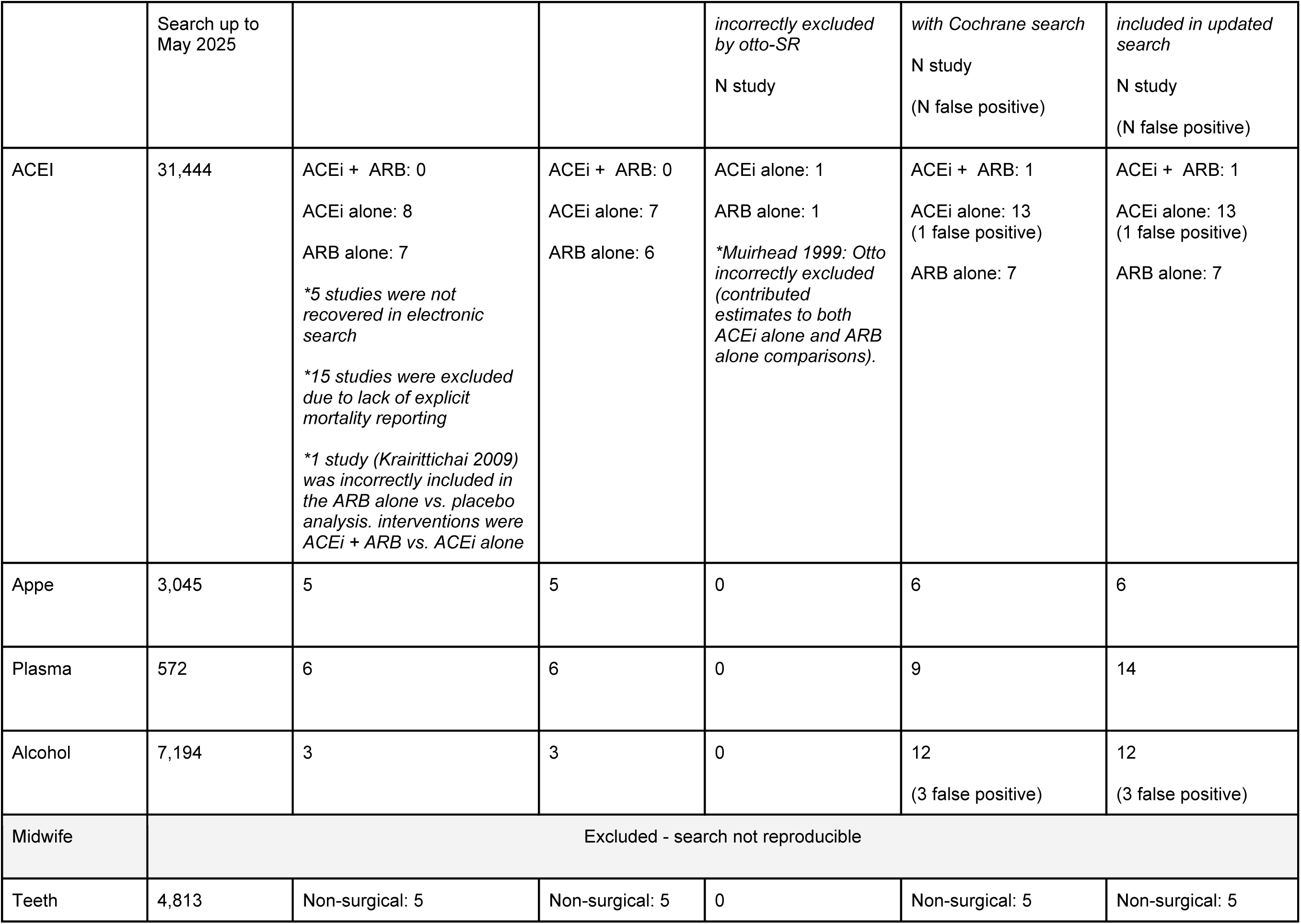
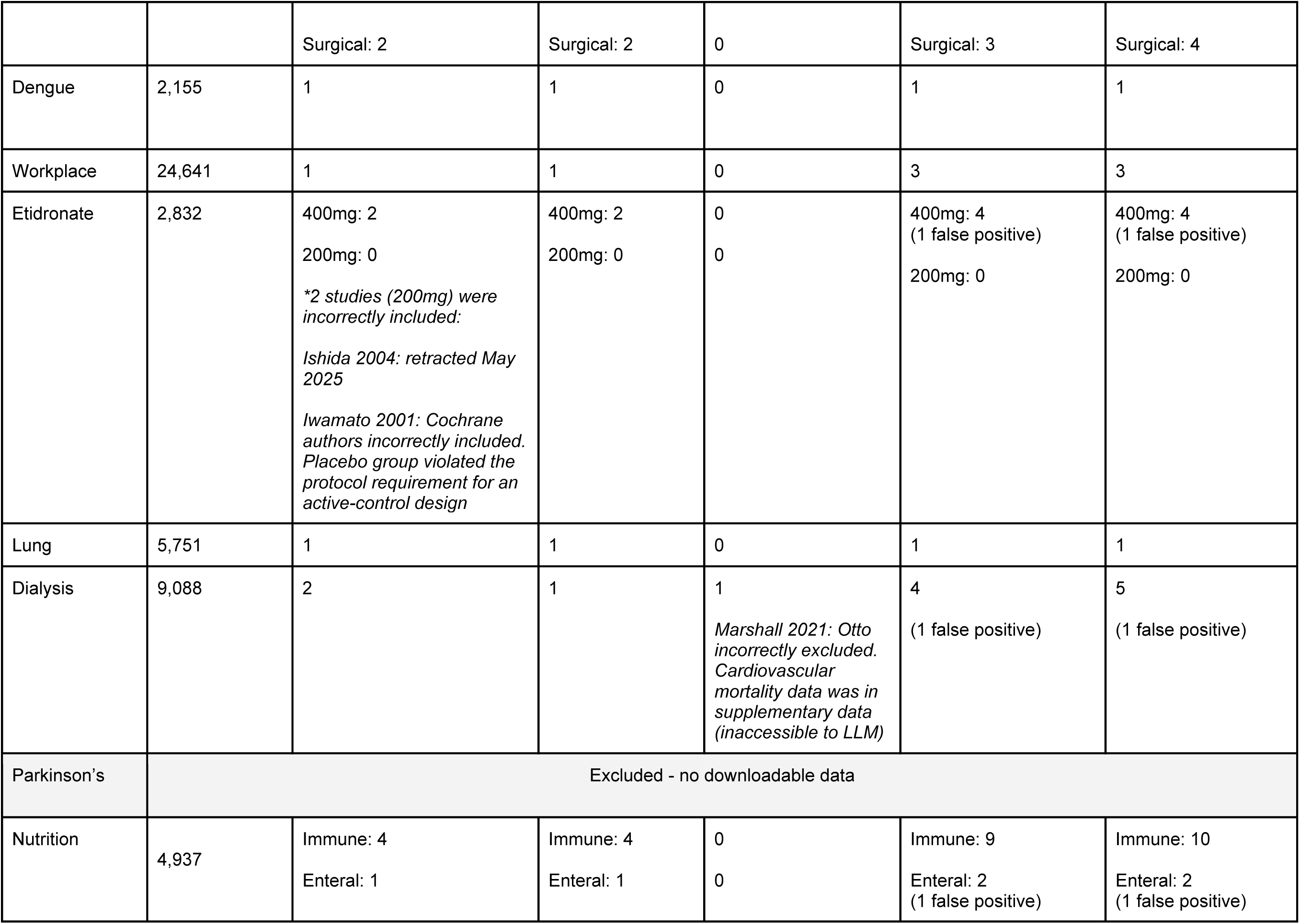
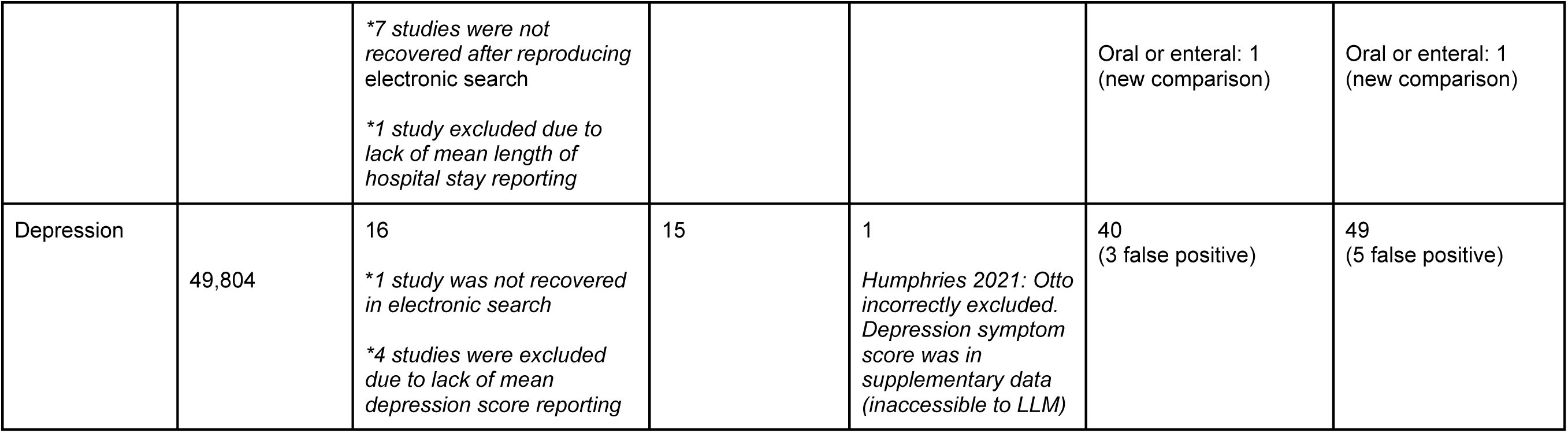
Phase 4: Descriptive Overview of Studies included and excluded in the Cochrane reproducibility analysis.

After filtering our results to match the original search date cutoffs, we identified 54 additional eligible studies (‘***Expanded***’) through *otto-SR* (median 2, IQR: 1 to 6.25 per review) that may have been missed in the original Cochrane reviews (Methods). *otto-SR* incorrectly included 10 false positive articles. However 9 of the 10 articles may have contained relevant data available through author data request.

Updating the search date to May 8, 2025 (‘***Update***’) identified another 14 new eligible studies (median 2.5, IQR 1 to 7.25 per review) (Table 9). The updated search identified two additional false positive studies, one of which may have contained relevant data through author data request.

Compared to the corrected per domain ROB2 assessments, *otto-SR* had high weighted accuracy (96.6%) and interrater concordance (0.96, Gwet AC2), with no major discrepancies (i.e., low vs. high) in overall RoB judgements (Supplementary Table 2, Supplementary Fig. 2,3)

In the ‘**Matched**’ comparison group, *otto-SR* produced meta-analyzed effect estimates that had overlapping 95% CIs with both the original Cochrane data and corrected datasets across all reviews (Fig 5b, left; Table 10). In the ‘**Expanded**’ analysis, two reviews (nutrition, depression) yielded new statistically significant effect estimates for the primary outcome (Fig 5b, middle), while the estimate from one review (alcohol) lost statistical significance compared to the original Cochrane estimates (Fig 5b, middle). These trends were consistent in the corrected ‘**Expanded**’, corrected ‘**Update**’, and *otto-SR* ‘updated’ analyses (Fig 5b, right).

**Table 10.**
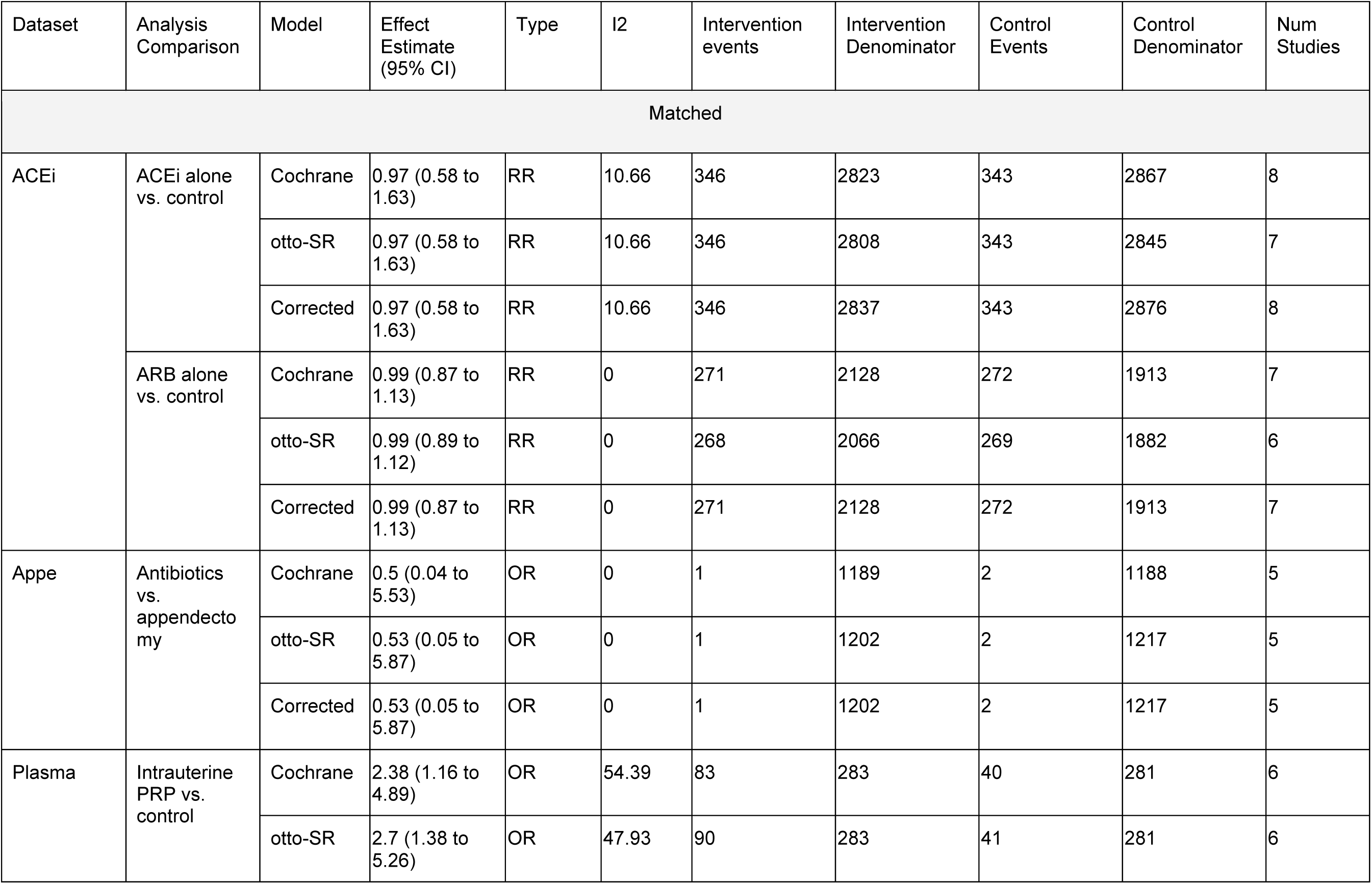
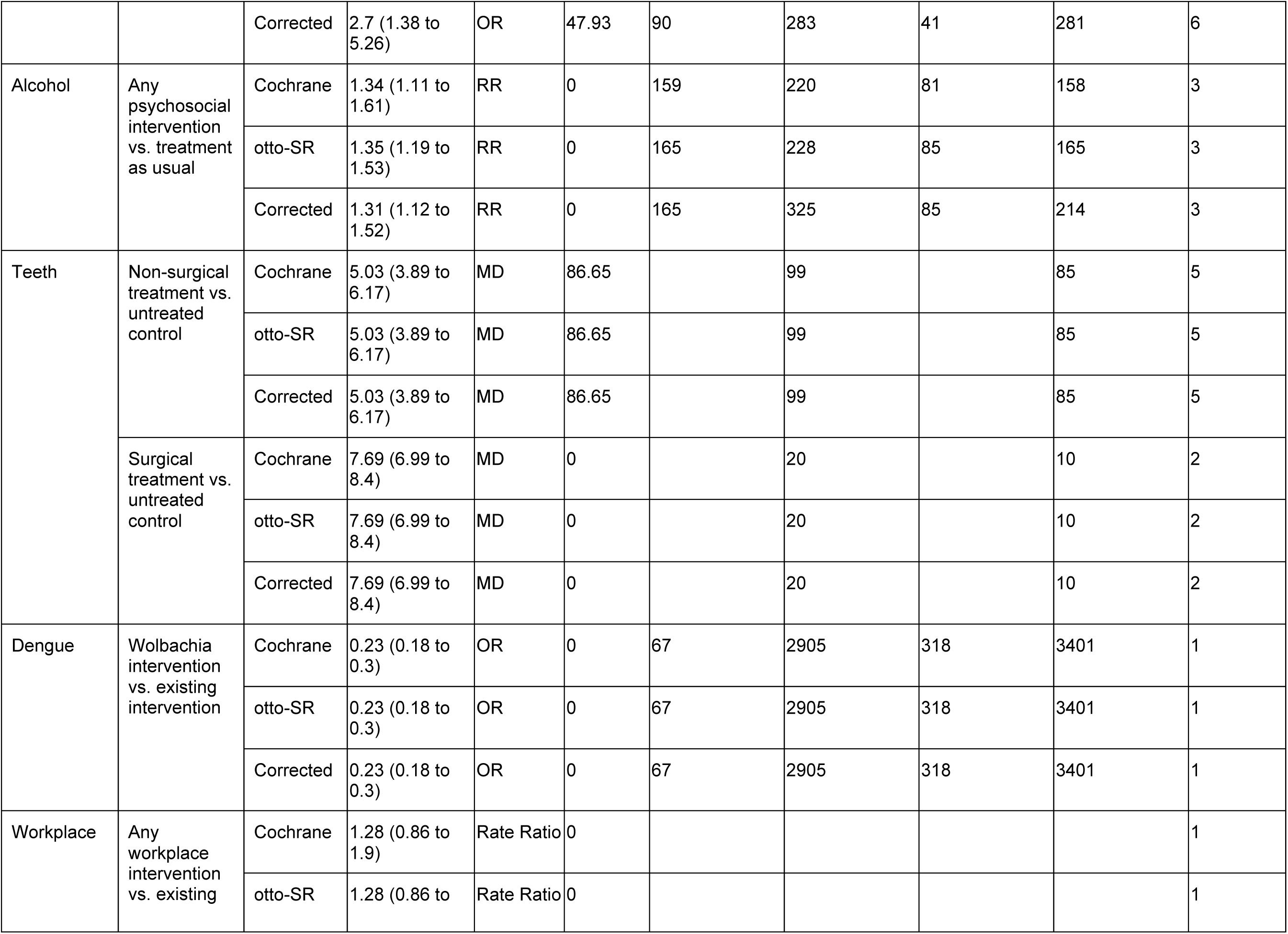
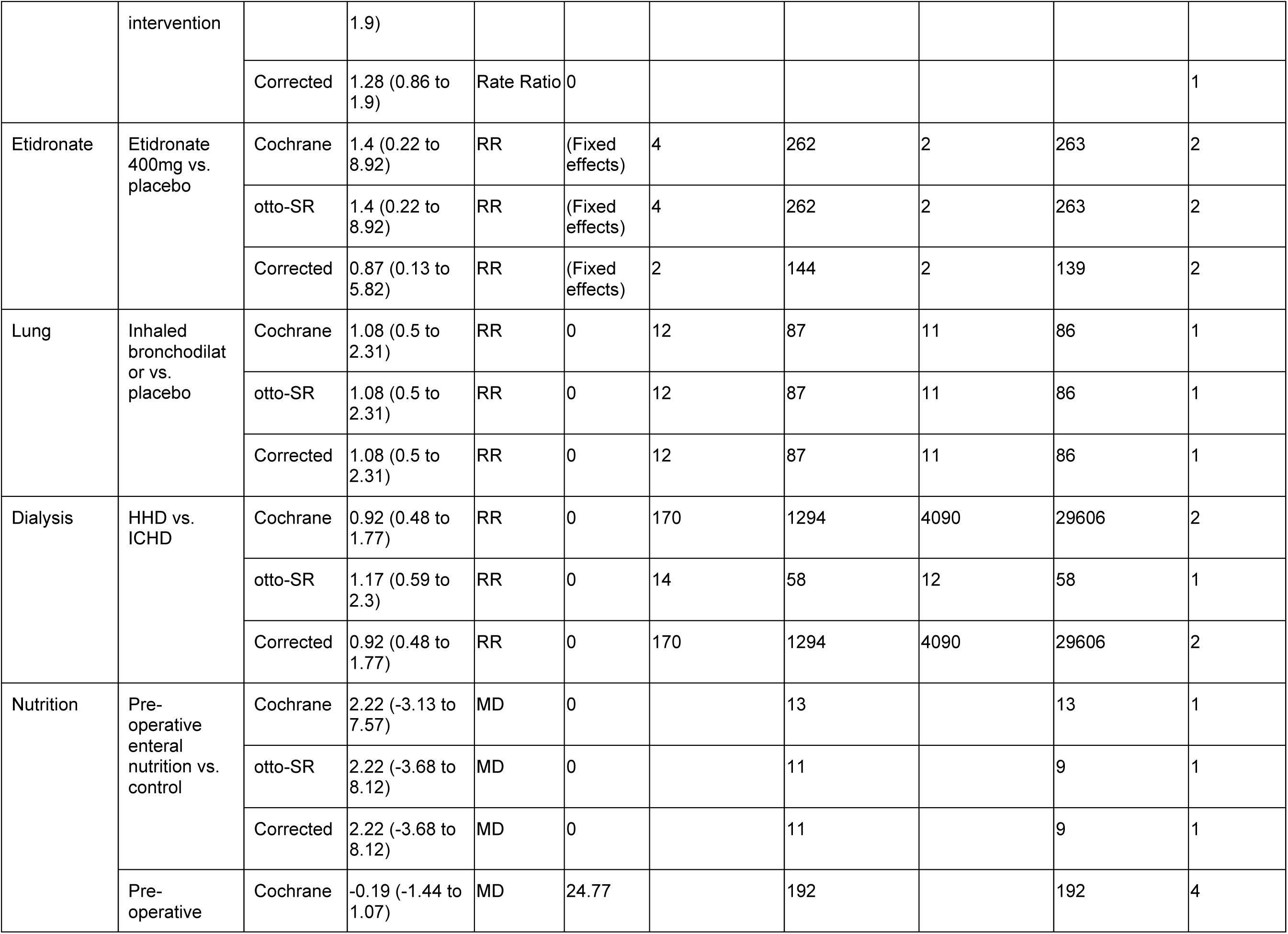
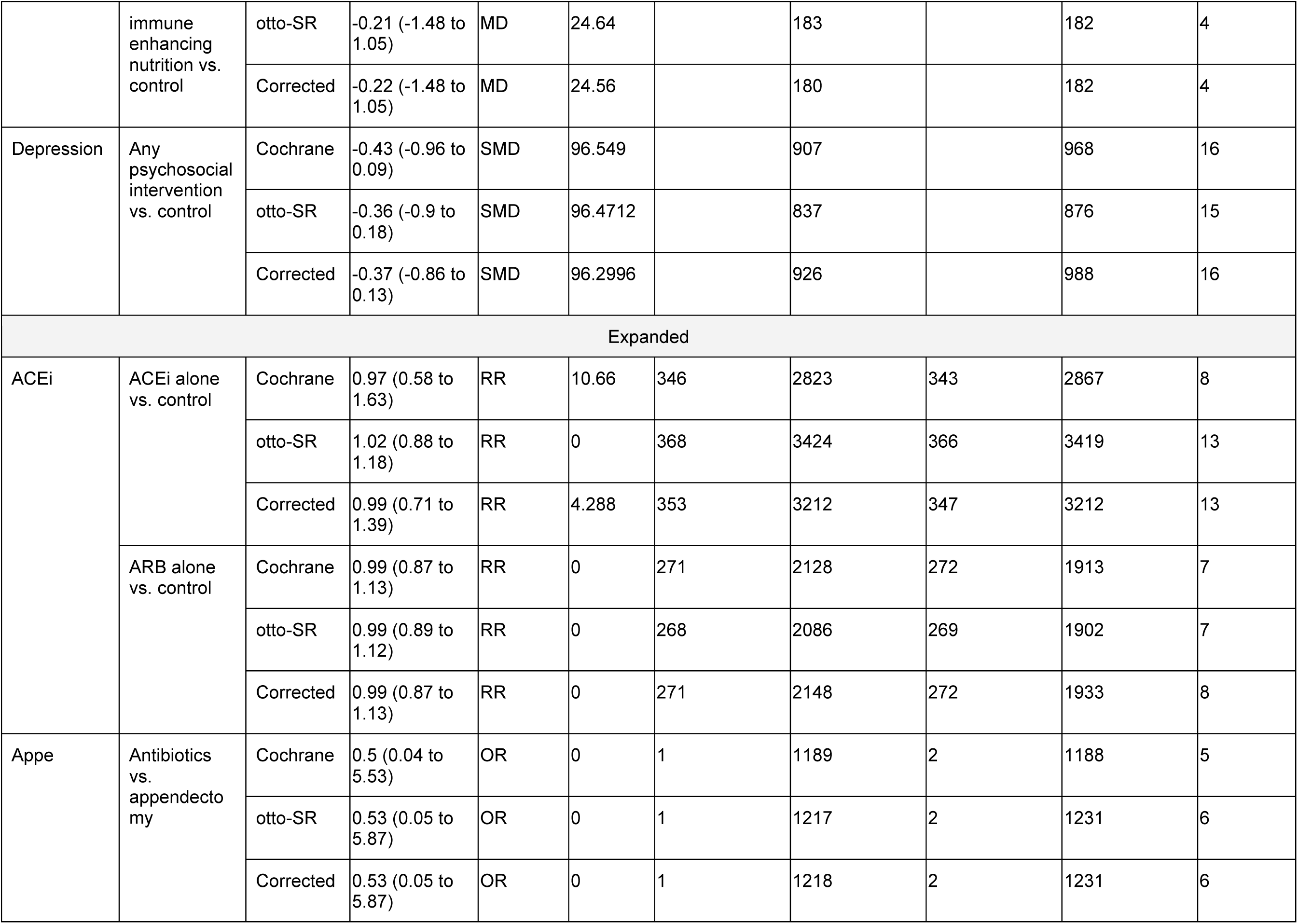
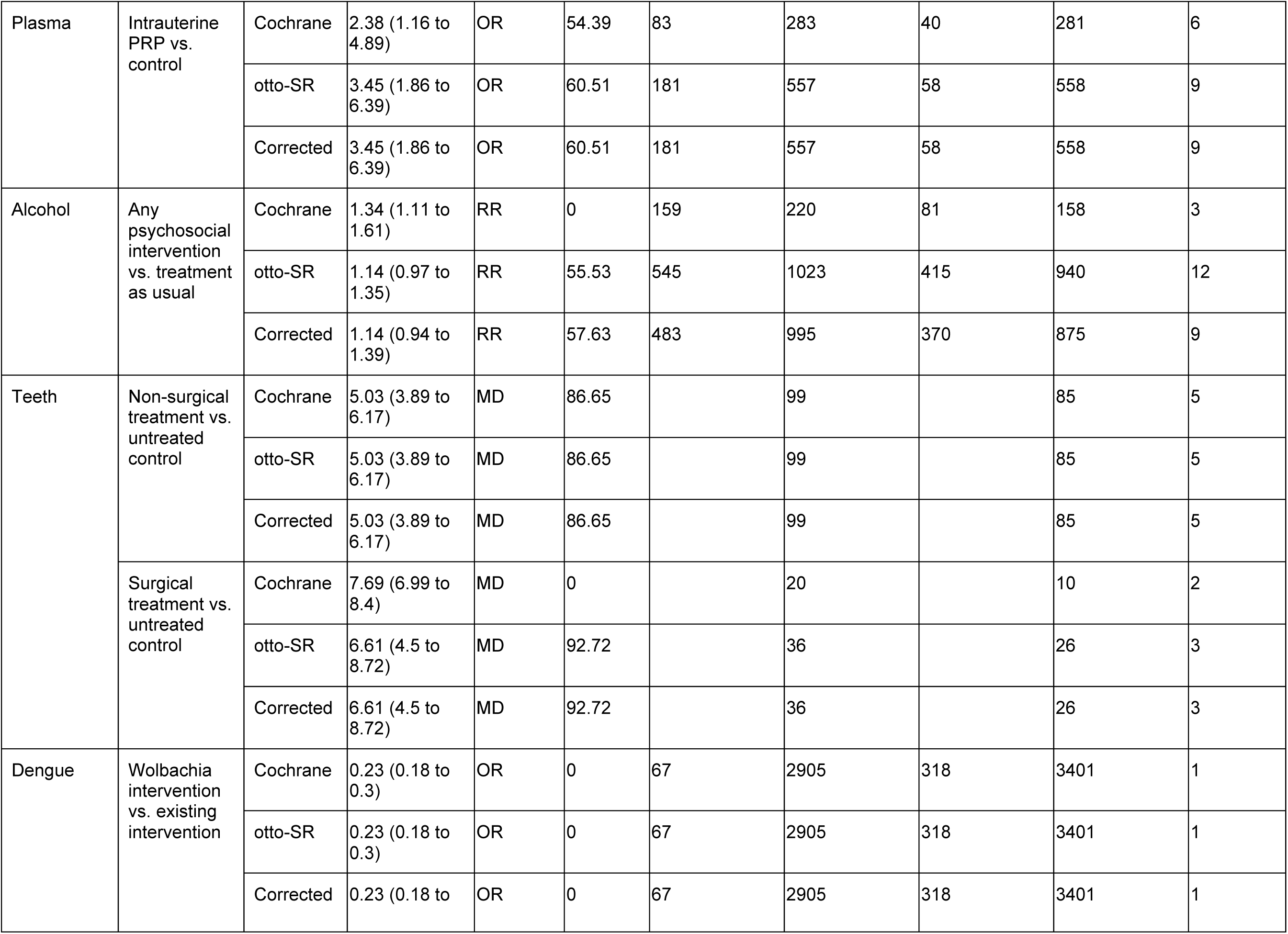
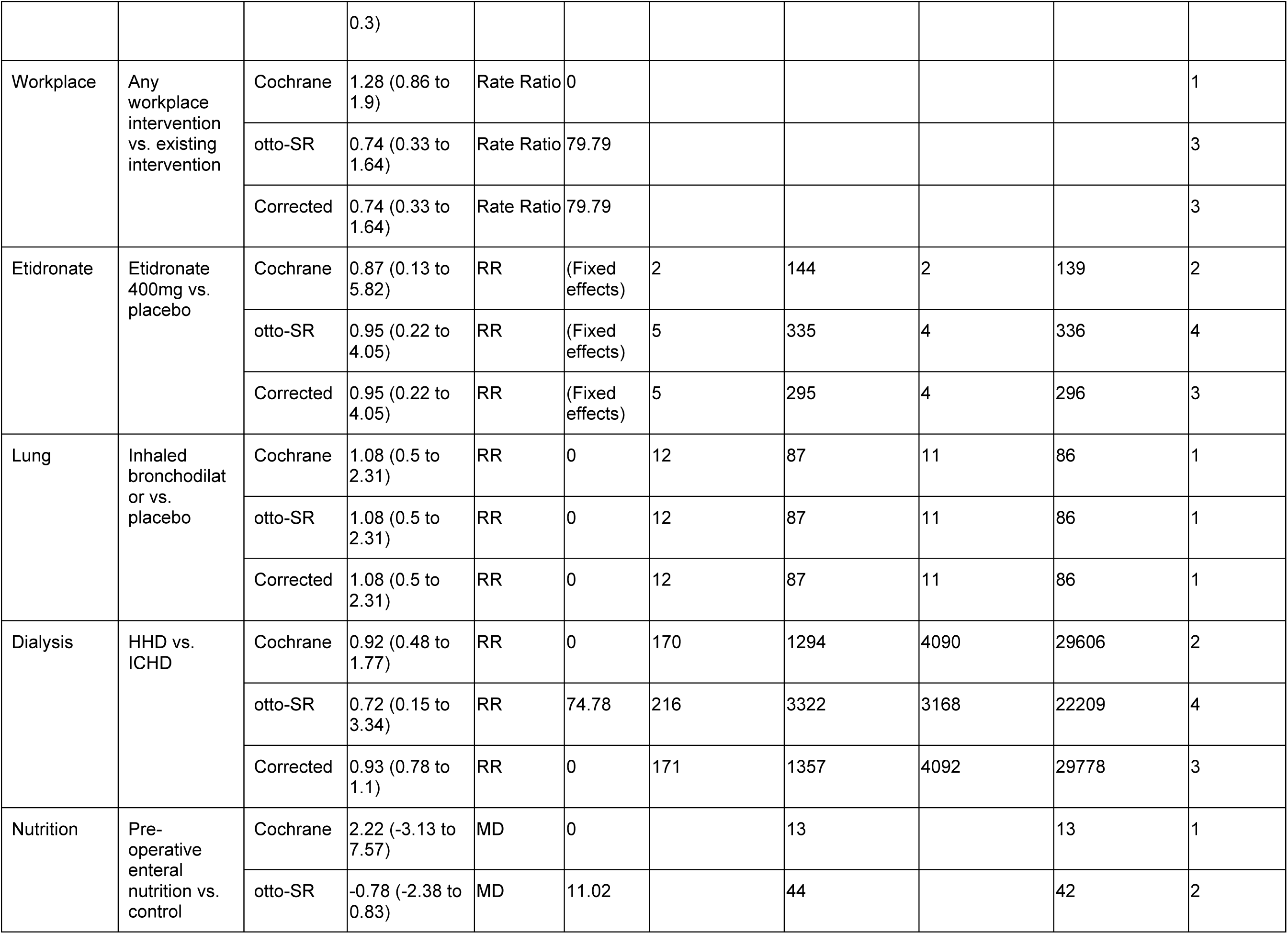
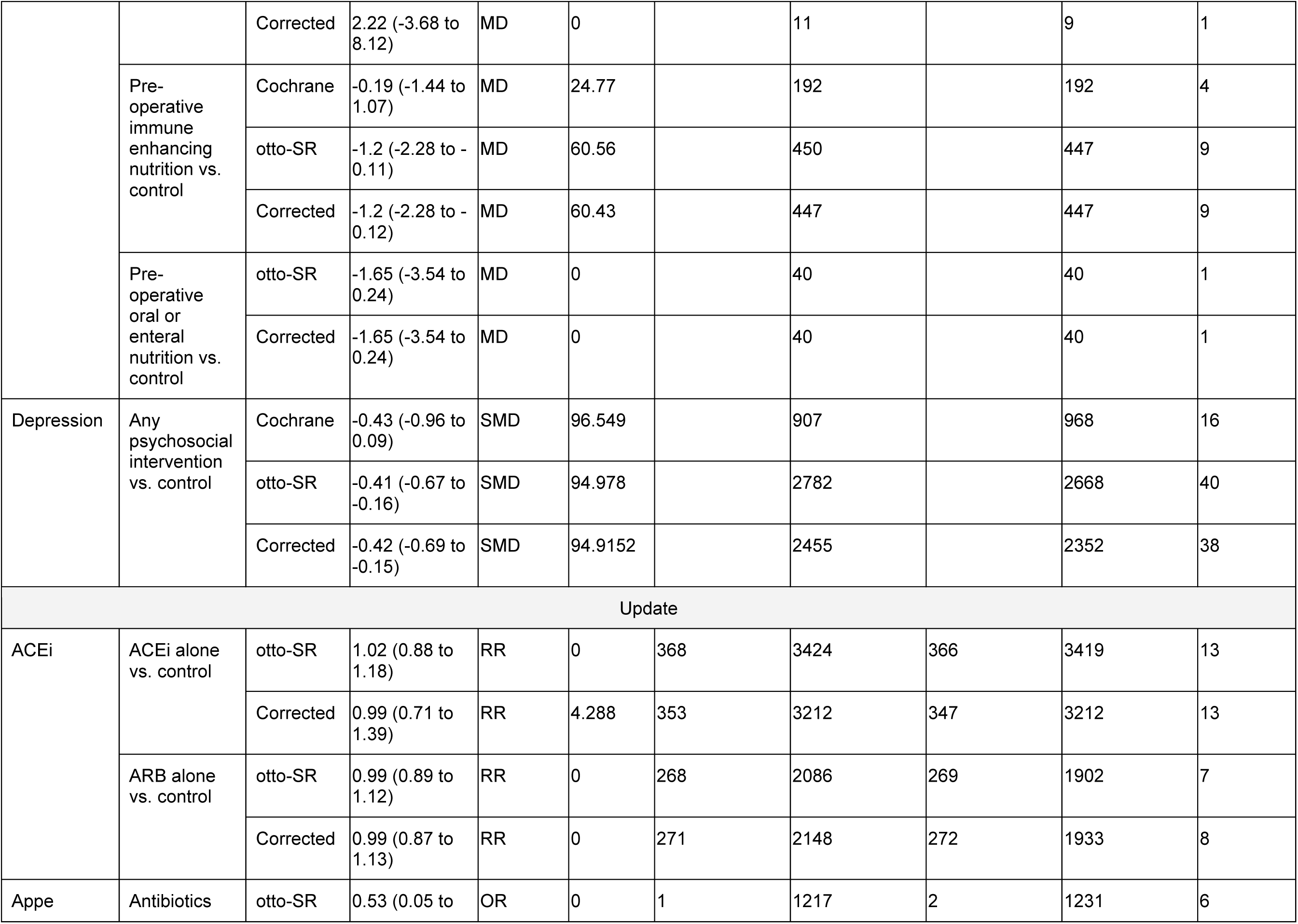
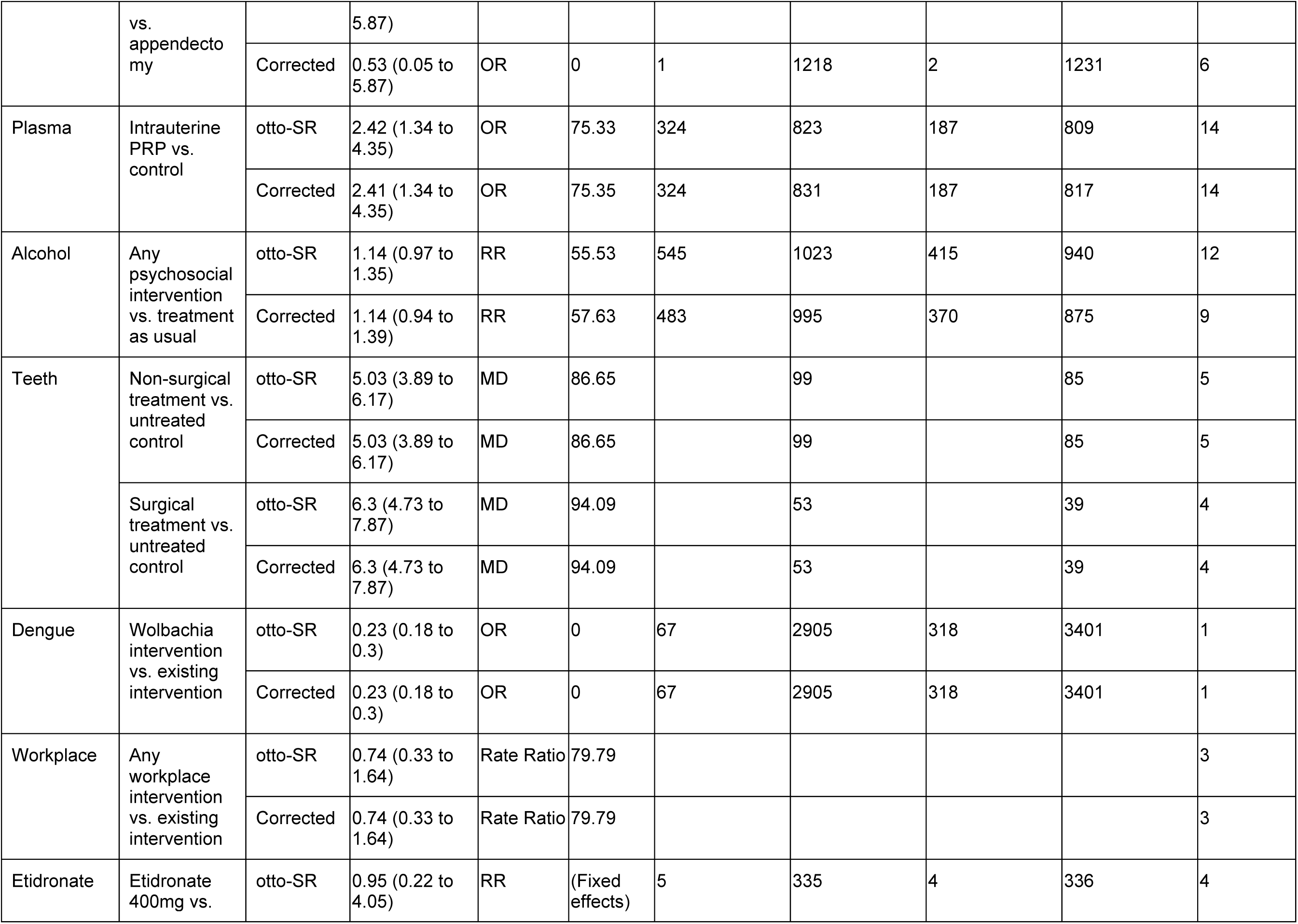
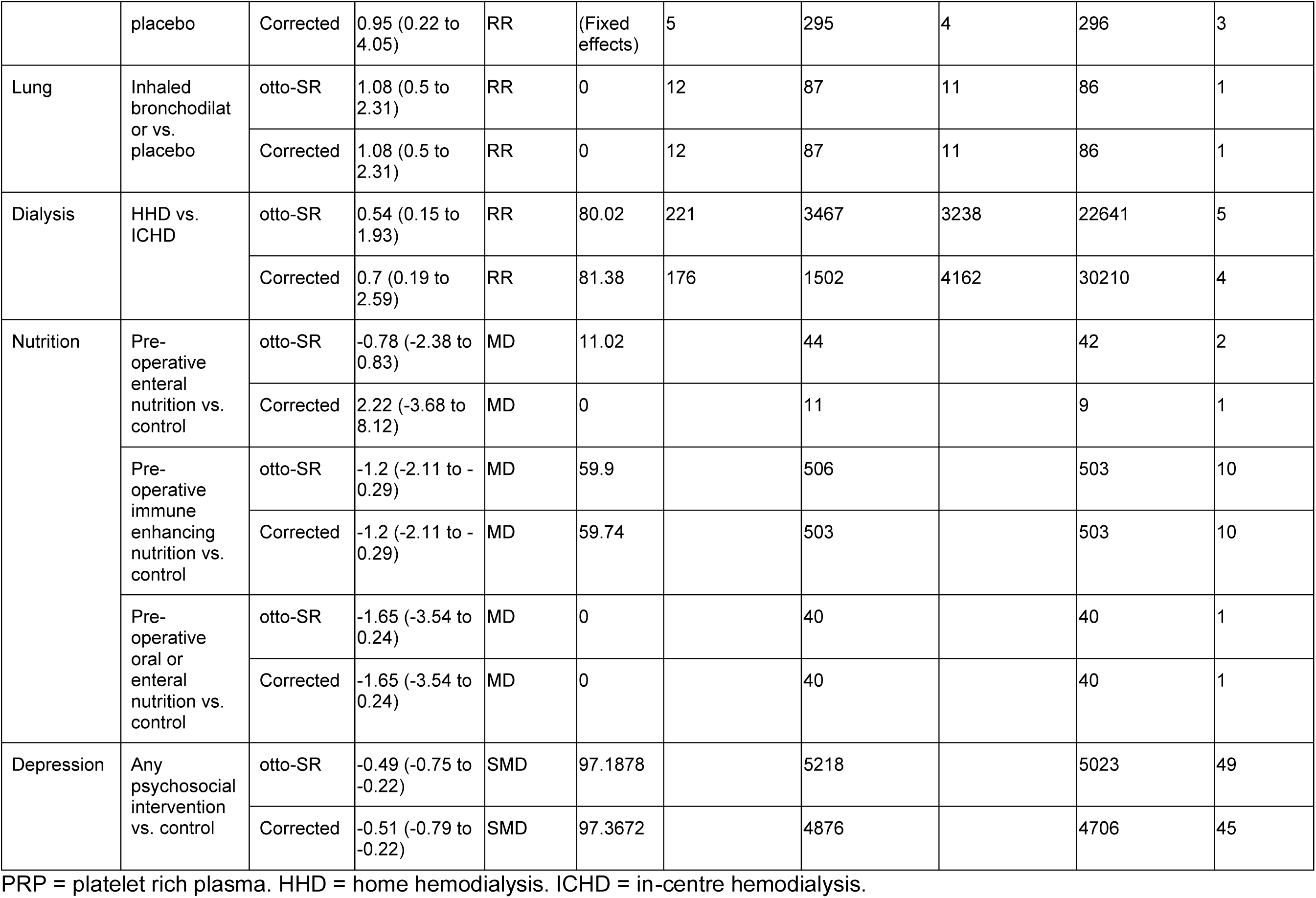
Phase 4: Meta-analyzed results between original cochrane authors, *otto-SR*, and corrected ground-truth across Cochrane reviews.

One illustrative example comes from the nutrition review, where *otto-SR* identified 5 additional studies in the ‘Expanded’ comparison. This led to the new finding that preoperative immune-enhancing supplementation before gastric surgery is associated with a significant one-day reduction in mean hospital stay compared to usual care (*otto-SR:* MD −1.20 [95% CI −2.28 to - 0.11], 9 studies vs Cochrane: MD −0.19 [−1.44 to 1.07], 4 studies). Detailed effect estimates and 95% CIs are provided for all groups and comparisons are provided in Table 10.

## Discussion

In this study, we demonstrate that *otto-SR*, an LLM-based SR automation pipeline, can accelerate key article screening, data extraction, and risk of bias assessment steps with high performance. We also demonstrate the feasibility of applying an LLM workflow to reproduce and update the primary analysis from multiple Cochrane reviews.

Our work has several strengths. First, we evaluated an LLM workflow for multiple phases of the SR process. Second, our phase 4 analysis examined downstream changes in effect estimates when reproducing and updating Cochrane reviews using automated screening, data extraction, and risk of bias assessment. Many prior studies have evaluated individual components in isolation, and do not assess how missed studies or extraction errors propagate into final meta-analyses.^[66–70]^ Third, otto-SR was applied using unaltered review objectives and protocols, demonstrating the potential for generalizable use of LLM-based workflows. This contrasts with prior work relying on review-specific prompt optimization,^[66,71]^ which may limit external applicability. Finally, we tested *otto-SR* on a diverse set of reviews including reviews of prevalence, prognosis, diagnostic test accuracy, and intervention effectiveness, thus increasing the generalizability of the findings.

Our work has several limitations. First, although our analysis was validated across a wide range of SRs, further research is needed to assess the generalizability to other clinical questions and qualitative reviews. Second, our LLM parser may have incorrectly extracted information from PDF articles. In future studies, enhanced vision capabilities of new models may better support screening/extraction efforts. Third, our workflow was limited to the main text of articles and did not extract data from supplementary tables or figures. While we could have manually incorporated these materials, we intentionally avoided human intervention to test the end-to-end capabilities of our automated workflow. Fourth, due to the vast number of data points (and their unstandardized nature), we employed an LLM-as-a-judge framework to assess data extraction accuracy. Although this approach has been previously validated,^[72,73]^ LLM judgements may still introduce errors. Fifth, we encountered instances where the original author’s decisions for data extraction appeared inaccurate. However, we addressed this by performing randomized and blinded adjudication, adopting best practices seen in radiology, ophthalmology, and clinical trials.^[32–35]^ This approach represented a methodological improvement over prior work that relied on self-created ground truths.^[9]^ Sixth, the GPT-4.1 knowledge cut-off was updated to June 2024 (released April 2025). However, our screening and data extraction benchmarks primarily involved closed-source data, and the April 2024 Cochrane reviews were closed access during the time of model pretraining (Cochrane articles are closed-access for a year after publication); mitigating concerns about potential pretraining data overlap. Finally, with respect to the Cochrane reproducibility assessment, it’s possible that content specific experts would have made different article inclusion and data extraction decisions. However, we closely adhered to each review’s protocol, contacted study authors to clarify methodological uncertainties, and documented all discrepancies.

Our findings highlight several opportunities for LLMs in SRs. First, workflows like *otto-SR* may be used to update existing SRs by leveraging their original published protocols. This provides a unique advantage: it enables validation of screening and extraction using the original review.

Second, the ability to rapidly screen articles, extract articles, and assess risk of bias would allow LLMs to support living SRs for which updates could be performed monthly, weekly, or even daily to ensure constant access to the best available cumulative evidence. This livingness reflects knowledge - it is always evolving and not fixed. Third, after further prospective validation, LLMs may be used to generate *de novo* reviews, provided that researchers develop clear research questions with detailed protocols akin to those registered in PROSPERO. In all cases, structured, transparent, and clear methodology is essential for ensuring interpretability, reproducibility, and high-quality automation.

The Phase 4 Cochrane reproducibility assessment identified common reproducibility challenges. All 12 reviews had issues with search reproducibility or methodological uncertainty (Table 8). These findings align with prior work by Rethlefsen et al.^[74]^ who found that only 1% of reviews have a fully reproducible search strategy. Previous studies have also shown that reproducibility failures can occur at every stage of the SR process.^[74–79]^ Furthermore, there are few SR reproducibility experiments; the results here (n=12 reviews) and by Hamilton et al.^[80]^ (n=10 reviews) are the largest to date. To address these challenges, published SRs should include: (i) a complete search strategy; (ii) raw search files (e.g., RIS file); (iii) raw data extraction outputs with data dictionaries; (iv) list of data procured via author correspondence; and (v) code used for analyses. Current reporting guidelines for systematic reviews (PRISMA) endorse most, but not all of these points.^[81]^ While Cochrane reviews routinely provide such materials, most other SRs do not, limiting reproducibility and external validation. Tools like *otto-SR* can support automatic generation and reporting of these files, reducing the burden on authors.

The high performance of LLMs in conducting evidence synthesis, as demonstrated in this study, may warrant consideration in how scientific content is published. While most research is written for human readers, the rise of LLM-supported evidence curation highlights the value of making studies machine-readable. Using structured formats like markdown or html (as now offered by Arxiv), and providing raw numerical data from figures could support this new paradigm.

### Conclusion

These findings represent an advancement in the development of SR automation tools. Downstream applications include rapid SR updates, truly ‘living’ reviews, and mass assessments of reproducibility across the SR literature. Future research should focus on developing comprehensive and complete benchmarks of SRs to better support and refine automation efforts, and evaluating the performance of LLMs on prospective de novo reviews. We also encourage research into the capabilities of LLMs for other SR workflow tasks, such as search term generation and statistical analysis. The implementation of more autonomously completed SRs could accelerate the synthesis of up-to-date evidence, saving thousands of hours of manual work, and transform how scientists, clinicians, and patients make decisions.

## Data Availability

All datasets and code used for data analysis will be made available on publication.

## Acknowledgements

We thank Guravneet Gill for assistance with database access; Zion Chan, Li Li, Lina Ghosn, and Isabelle Boutron for discussions during project development; and Luke Son for technical insights. We acknowledge all systematic review authors whose published data and methodologies enabled this reproducibility assessment.

## Author Information

These authors contributed equally: C.C, R.A., P.C. Author contribution statements: C.C, R.A, N.B contributed to the conception and design of the work. C.C, R.A, K.M, M.C, E.F, A.S, R.K, R.S, D.M, J.L, J.J, D.C, J.G, S.L contributed to data acquisition, cleaning, human comparisons, and human arbitration. C.C, R.A, P.C, J.S, S.J, J.X, K.Z generated code for evaluations and benchmarking. C.C, R.A, P.C, L.X.G analyzed study data. N.B, G.M.C, D.M, A.T, D.B.E, R.K.A, L.H, J.P, A.S.D, M.N, A.A.L, B.T, M.W, H.W contributed to project supervision and provided feedback on the study. C.C, R.A, P.C, prepared the original draft of the manuscript with input from all co-authors. All authors were responsible for review and editing of the manuscript. All authors debated, discussed, edited, and approved the final version of the manuscript.

## Funding and Ethics Declaration

There was no direct funding support for this manuscript.

N.B reports grants from the Public Health Agency of Canada through Canada’s COVID-19 Immunity Task Force, the World Health Organization Health Emergencies Programme, the Robert Koch Institute, the Canadian Medical Association Joule Innovation Fund, the Canadian Association of Emergency Physicians and Alberta Health Services Emergency Strategic Clinical Network. Disclosures for G.M.C. can be found at http://arep.med.harvard.edu/gmc/tech.html.

R.K.A. is employed at OpenAI and owns stock as part of the standard compensation package.

R.A. reports grants from the CIHR Institute of Genetics.

A. C. P.C. J.S. are founders of and hold equity in Otto Science Institute. N.B, D.M, A.C.T, A.B hold equity in Otto Science Institute. Of note, Otto Science institute was founded after the time of initial manuscript writing.

No funding source had any role in the design of this study, its execution, analyses, interpretation of the data, or decision to submit results.

## Code and Dataset Availability

Datasets will be made available upon reasonable request.

Code used for data analysis and figure generation will be made available on publication. The OttoSR platform is commercially available at ottosr.com (contact). Source code is proprietary and not openly distributed. To support reproducibility, researchers can reproduce all reported results by running the same versioned otto-SR workflow via access at ottosr.com (contact).

---

**Supplementary figure 1.**
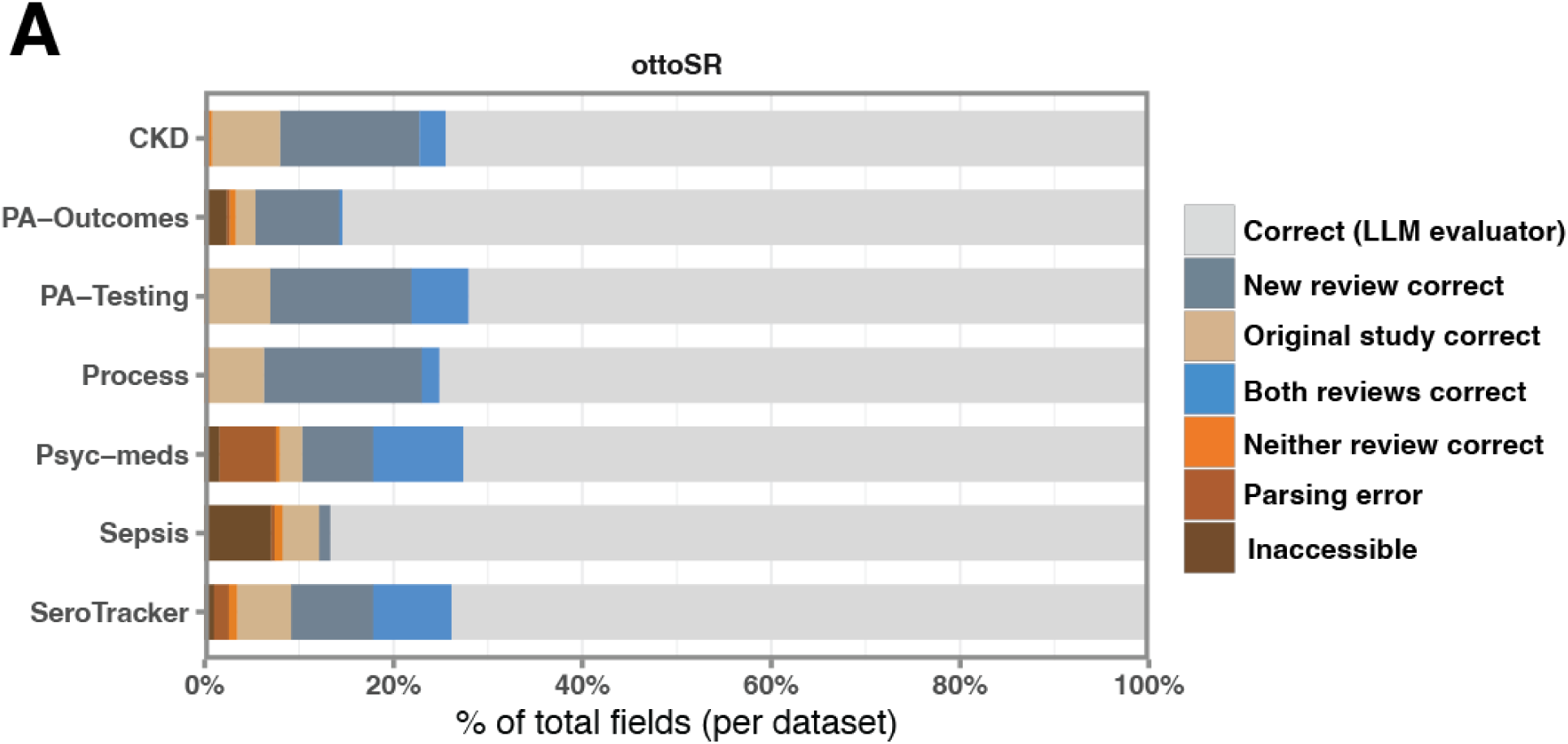
Data extraction answer classification. Bar graph displaying answer classification resulting from dual human adjudication across all 7 evaluated systematic reviews.

**Supplementary figure 2.**
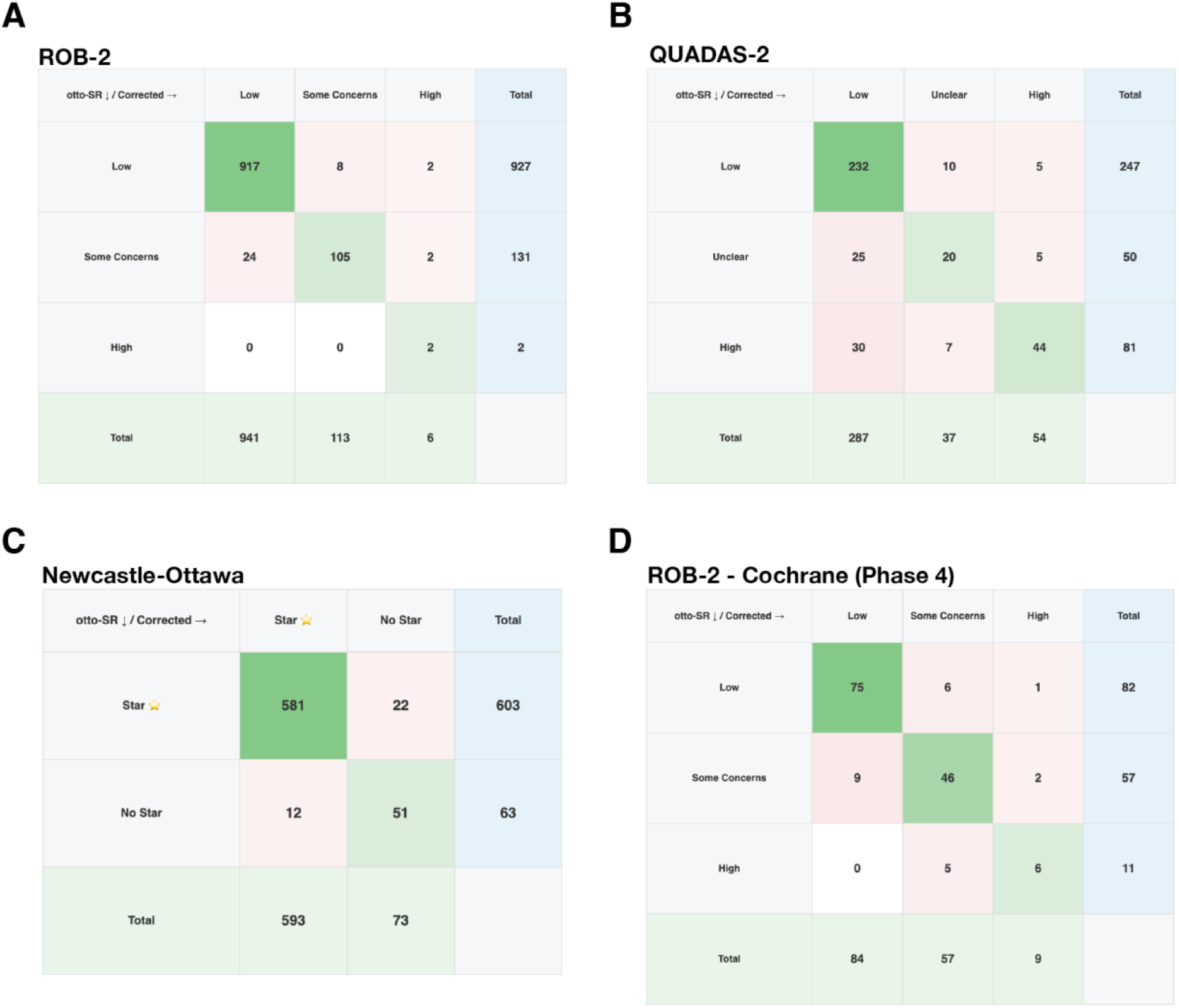
Confusion matrix of per domain risk of bias assessments. Confusion matrix displaying per domain risk of bias assessments from the otto-SR RoB agent relative to original review risk of bias assessment decisions for (A) ROB2, (B) QUADAS-2, (C) Newcastle-Ottawa, and (D) ROB2 in the phase 4 Cochrane reproduction; relative to a corrected reference standard.

**Supplementary figure 3.**
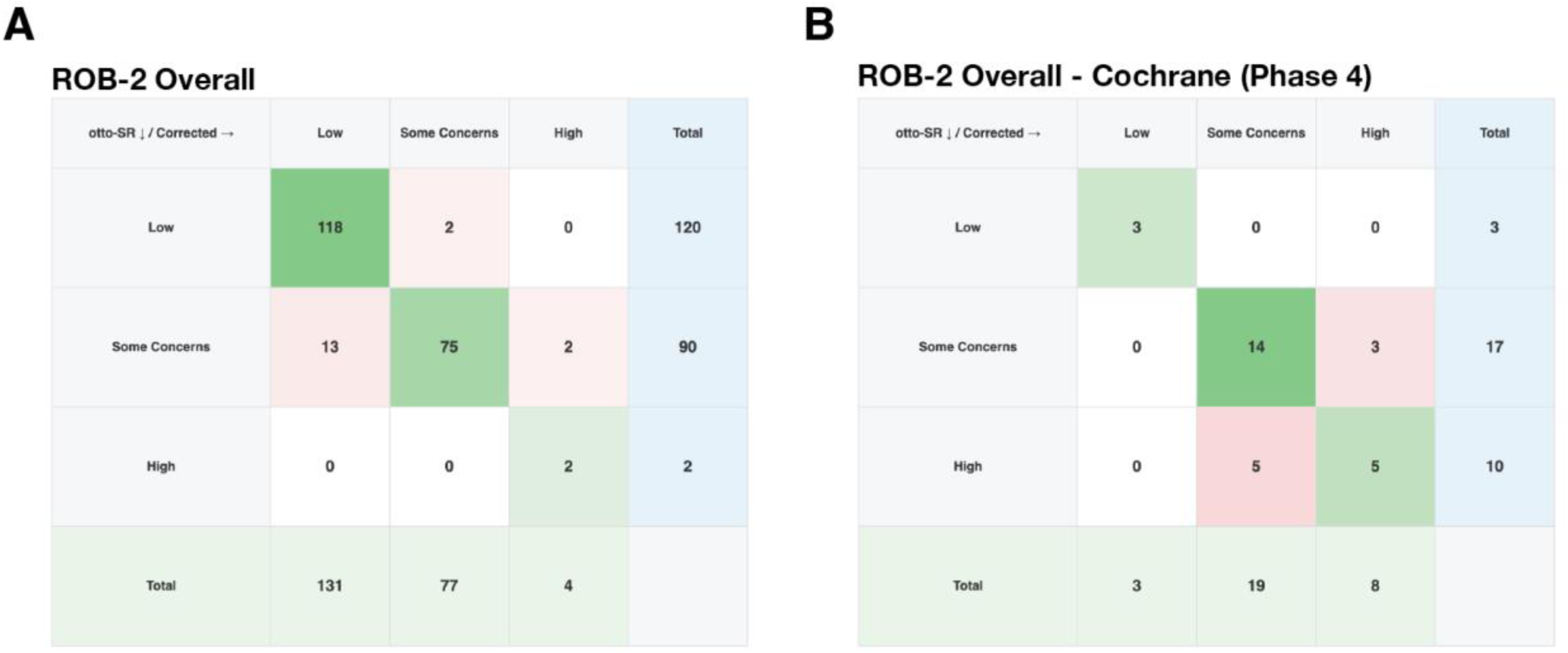
Confusion matrix of overall RoB assessments. Confusion matrix displaying overall RoB risk of bias assessments from the otto-SR RoB agent and original review risk of bias assessment decisions, relative to a corrected reference standard for (A) Phase 3 ROB2, and (B) Phase 4 Cochrane reproducibility. Matrices were only generated for the ROB2 tool, as other tools only provided assessments on a per domain/question basis.

